# Multi-ancestry GWAS of diarrhea during acute SARS-CoV2 infection identifies multiple novel loci and contrasting etiological roles of irritable bowel syndrome subtypes

**DOI:** 10.1101/2024.04.03.24305274

**Authors:** Ninad S. Chaudhary, Catherine H. Weldon, Priyanka Nandakumar, Janie F. Shelton, 23andMe Research Team, Michael V. Holmes, Stella Aslibekyan

## Abstract

A substantial proportion of acute SARS-CoV2 infection cases exhibit gastrointestinal symptoms, yet the genetic determinants of these extrapulmonary manifestations are poorly understood. Using survey data from 239,866 individuals who tested positively for SARS-CoV2, we conducted a multi-ancestry GWAS of 80,289 cases of diarrhea occurring during acute COVID-19 infection (33.5%). Six loci (*CYP7A1, LZFTL1- -CCR9, TEME182, NALCN, LFNG, GCKR*) met genome-wide significance in a trans-ancestral analysis. The top significant GWAS hit mapped to the *CYP7A1* locus, which plays an etiologic role in bile acid metabolism and is in high LD (r^2^= 0.93) with the *SDCBP* gene, which was previously implicated in antigen processing and presentation in the COVID-19 context. Another association was observed with variants in the *LZTFL1–CCR9* region, which is a known locus for COVID-19 susceptibility and severity. PheWAS showed a shared association across three of the six SNPs with irritable bowel syndrome (IBS) and its subtypes. Mendelian randomization showed that genetic liability to IBS-diarrhea increased (OR=1.40,95%,CI[1.33-1.47]), and liability to IBS-constipation decreased (OR=0.86, 95%CI[0.79-0.94]) the relative odds of experiencing COVID-19+ diarrhea. Our genetic findings provide etiological insights into the extrapulmonary manifestations of acute SARS-CoV2 infection.

## INTRODUCTION

Diarrhea is a common extrapulmonary gastrointestinal (GI) symptom of acute COVID-19. Retrospective studies estimate that the prevalence of COVID-19 related diarrhea (defined herein as COVID-19+ diarrhea) varies between 7% and 18% for population based and hospital based cohorts, respectively ^1–3^. There is growing evidence that SARS-CoV2 tropism (the ability of a virus to infect multiple cell types) in gastrointestinal tissues may lead to alteration of gut microbiota ^4,5^ and persistence of virus in the gastrointestinal system, which is associated with a higher risk of post-acute sequelae such as long COVID^6^. SARS-CoV2 RNA can be detected in fecal samples of COVID-19+ patients up to 4 months after acute infection, with fecal viral RNA detectable for longer durations among individuals with COVID-19+ diarrhea as compared to those without diarrhea^7^. Yet, the gastrointestinal symptoms of SARS-CoV2 positive individuals have not been studied in GWAS.

The SARS-CoV virus enters the body via angiotensin-converting enzyme 2 (ACE2) and co-receptor transmembrane protease serine 2 (TMPRSS2)^8^ receptors that are predominantly expressed in the lung. ACE2 and TMPRSS2 receptors are also expressed in gastric mucosal cells, enterocytes, and colonocytes^9^, making the GI tract a potential extrapulmonary site of SARS-CoV2 infection. SARS-CoV2 RNA and viral proteins were detected in epithelial cells from intestinal biopsies of COVID-19 patients with acute infection^10^. In postmortem data from 13 patients, SARS-CoV2 subgenomic RNA was detected in the small intestinal tissues from eight patients, indicative of viral replication^11^. It is not clear whether GI symptoms are the direct consequence of SARS-CoV2 infection of intestinal cells or due to a systemic immune-inflammatory response. GWAS can provide insights into the genetic architecture of disease and shed light on potential etiological mechanisms. To date, no GWAS studies of COVID-19+ diarrhea have been conducted. We sought to bridge this gap by conducting a GWAS of self-reported diarrhea symptoms among 239,866 COVID-19 test positive 23ndMe research participants.

## RESULTS

Of the individuals who self-reported testing positive for SARS-CoV2, 33% reported diarrhea as a symptom (N=80,289 cases of COVID-19+ diarrhea / 239,866 acute COVID-19 cases who tested positive). Among 80,289 COVID-19+ diarrhea cases, the vast majority reported diarrhea symptoms at baseline (N=79,361 cases) with a few individuals reporting diarrhea at 1 month (N=313 cases), 2 months (N=346 cases),or 3 months of follow up (N=269 cases). Women accounted for the majority of the survey participants (71.1% of COVID-19+ diarrhea cases and 65.4% of controls), and were more likely to experience COVID-19+ diarrhea than men (adjusted OR[95%CI] =1.29[1.27,1.31]). Compared to those with COVID-19+ who didn’t experience diarrhea, individuals with COVID-19+ diarrhea were younger, more likely to be of non-European ancestry, and to have diabetes, depression, and high triglyceride levels (**Table 1**).

**Table 1.** Characteristics of 239,866 research participants in 23andMe COVID-19+ study.

| Characteristics | COVID-19+<br>diarrhea | COVID-19+<br>without diarrhea |
| --- | --- | --- |
| No | 80,289 | 159,577 |
| **Demographics** |  |  |
| Age mean (SD) | 43.23 (14.12) | 44.11 (15.10) |
| Female N (%) | 57,065 ( 71.1) | 104,329 (65.4) |
| Education in years mean (SD) | 15.06 (2.37) | 15.34 (2.47) |
| Ancestry N (%) |  |  |
| *African American* | 3297 ( 4.1) | 7179 ( 4.5) |
| *East Asian* | 1282 ( 1.6) | 3047 ( 1.9) |
| *European* | 59821 ( 74.5) | 120470 (75.5) |
| *Latinx* | 15508 ( 19.3) | 27656 (17.3) |
| *South Asian* | 381 ( 0.5) | 1225 ( 0.8) |
| BMI mean (SD) | 30.05 (7.55) | 28.55 (6.82) |
| Tobacco Use N (%) | 30,091 ( 38.5) | 53,654 (34.5) |
| Two or more alcohol drinks per week –<br>current N (%) | 6,398(23.0) | 33,544(23.2) |
| **Health Comorbidities** |  |  |
| High blood pressure N (%) | 20,363 ( 25.8) | 34,836 (22.2) |
| Depression N (%) | 32,418 ( 44.5) | 50,773 (34.9) |
| Diabetes N (%) | 10,477 ( 14.1) | 16,083 (10.9) |
| High total cholesterol N (%) | 5679 ( 19.1) | 10371 (16.8) |
| High triglycerides N (%) | 4277 ( 26.3) | 7194 (21.1) |
| **COVID-19+ related characteristics** |  |  |
| COVID-19+ and hospitalized N (%) | 5,111 ( 6.6) | 4,570 ( 3.0) |

| Characteristics | COVID-19+<br>diarrhea | COVID-19+<br>without diarrhea |
| --- | --- | --- |
| **COVID-19+ and severe respiratory disease**<br>**N (%)** | 8,170 ( 10.2) | 7,702 ( 4.8) |

SD=standard deviation, IQR=Interquartile-range, COPD=Chronic Obstructive Pulmonary Disease, BMI=Body-Mass Index, Severe respiratory disease= pneumonia, difficulty breathing that may have required supplementary oxygen, or ventilatory support

Individuals of East Asian (adjusted OR[95%CI]=0.85[0.79,0.94]) or African American (adjusted OR [95%CI]=0.92[0.88,96]) ancestry had lower relative odds of reporting diarrhea compared to those of European ancestry, whereas Latinx individuals were at higher relative odds of reporting diarrhea (adjusted OR[95%CI]=1.21[1.09,1.14]). Those reporting COVID-19+ diarrhea were more than twice as likely to be hospitalized during the acute infection (6.6% in COVID-19+ diarrhea cases vs 3.0% in COVID-19+ controls; corresponding to an adjusted relative odds of 2.56; 95%CI: 2.46, 2.57) and were more likely to have severe respiratory disease (10.2% in COVID-19+ diarrhea cases vs 4.8% in COVID-19+ diarrhea controls; adjusted relative odds of 2.38; 95%CI: 2.30, 2.46) after adjusting for age, sex, and ancestry. Furthermore, those with COVID-19+ diarrhea had twice the odds of subsequently experiencing long COVID (43% in COVID-19+ diarrhea cases vs 23% in COVID-19+ diarrhea controls; adjusted relative odds of 2.53; 95%CI: 2.46, 2.61) and of long COVID impacting daily living activities (16% of COVID-19+ diarrhea cases vs 9.6% of COVID-19+ diarrhea controls; adjusted relative odds of 2.27; 95%CI: 2.21, 2.34).

### GWAS

We conducted GWAS within each of European, African American, Latinx, East Asian, and South Asian ancestry groups, and then meta-analyzed across ancestries using fixed effect modeling. Individuals of European ancestry contributed 75% of the sample size to multi-ancestry analysis, with African-Americans contributing 4%, Latinx 18%, East Asians 2%, and South Asians 1%. We identified six distinct loci associated with COVID-19+ diarrhea in multi-ancestry analysis (**Table 2**, **Figure 1**). The associations of index variants were statistically significant at baseline but not at the following time points (1 and 3 months after onset of acute infection) likely owing to lack of statistical power (**Supplementary Table 1**). The top statistically significant index variant at chr8q12.1 (rs10504255) is situated in the intergenic region of genes *UBXN2B* and *CYP7A1,* located 4kb upstream of the *CYP7A1* gene. rs10504255 (A/G with G being the effect allele) was associated with COVID-19+ diarrhea with OR[95% CI] = 0.94[0.92,0.95], p= 2.6×10^-16^. The credible set from multi-ancestry analysis contained 16 variants covering a 100.4-kilobase(kb) region (**Supplementary Figure 1**). The association was primarily driven by individuals of European ancestry (p = 1.08 × 10^-14^) as compared to other ancestries (**Supplementary Table 2**). In the analysis of expression quantitative trait loci (eQTL), rs10504255 was found to be in high LD with a variant (rs9297994) associated with *CYP7A1* expression in the thyroid gland (p = 8.1×10^-9^) and a variant (rs8192870) associated with expression of a nearby gene (*SDCBP*, r^2^=0.94, p=9.9×10^-10^) in left ventricular myocardium (**Supplementary Table 3**)^12^. Previous studies have shown *SDCBP* expression to correlate with *HLA-DPB1* expression in normal lung tissue ^13^. The variant rs35044562 (alleles A/G with G being the effect allele) on chr3p21.31 lies in the intergenic region of *LZTFL1* and *CCR9* (**Supplementary Figure 2**). *LZTFL1* is widely expressed in ciliated epithelial cells in lungs ^14^. Additional genes in this regulatory locus include *CCR9,* primarily expressed in immune cells, and *SLC6A20* in the gastrointestinal tract.

**Figure 1:**
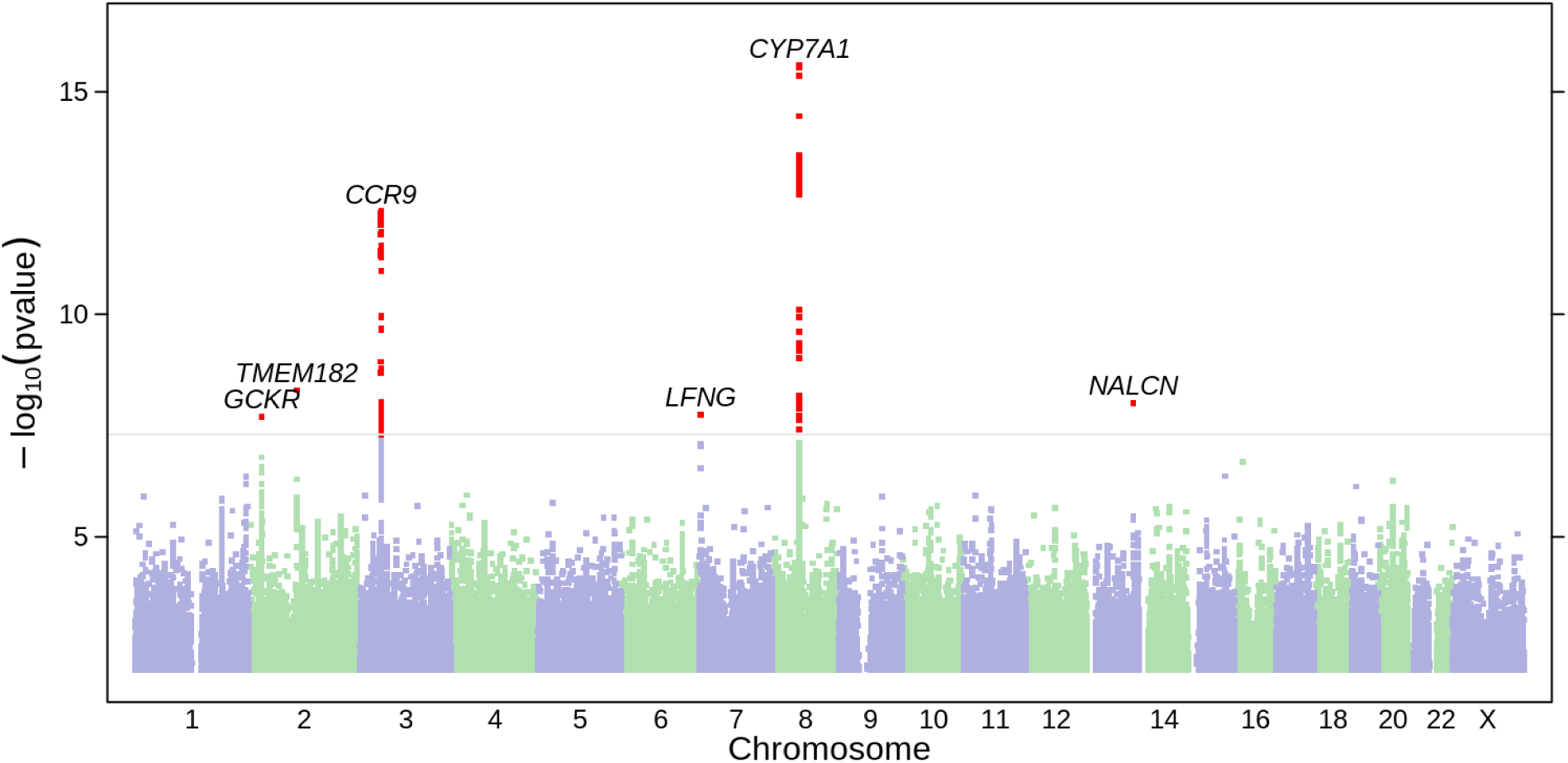
Manhattan plot of COVID-19+ diarrhea among 23andMe participants who tested positive for SARS-CoV2. Manhattan plot depicts findings from meta-analysis of five ancestral groups (European, African, Latinx, East Asian, and South Asian). X-axis represents chromosomal position for each SNP. Y-axis represents negative log p values based on logistic regression model under the additive model. Statistically significant variants are highlighted in red. The regions of associations are annotated with index variants.

**Table 2:** Statistically significant genetic variants associated with COVID-19+ diarrhea on meta-analysis.

| SNP | Chr | Position | Alleles | Gene | Effect allele | EAF | OR(95%CI) | P value |
| --- | --- | --- | --- | --- | --- | --- | --- | --- |
| rs10504255 | chr8 | 58485902 | A/G | UBXN2B--[]-CYP7A1 | G | 0.344 | 0.94(0.92,0.95) | 2.56X10<sup>-16</sup> |
| rs35044562 | chr3 | 45867532 | A/G | LZTFL1--[]--CCR9 | G | 0.078 | 1.10(1.07,1.13) | 4.88X10<sup>-13</sup> |
| rs75683620 | chr2 | 103792340 | A/G | TMEM182---[] | G | 0.9991 | 0.67(0.59,0.77) | 5.04X10<sup>-09</sup> |
| rs536843010 | chr13 | 101100662 | A/C | [NALCN] | C | 0.991 | 0.81(0.75,0.87) | 9.82X10<sup>-09</sup> |
| rs13245319 | chr7 | 2514631 | C/T | GRIFIN--[]-LFNG | T | 0.021 | 1.16(1.10,1.22) | 1.79X10<sup>-08</sup> |
| rs1260326 | chr2 | 27508073 | C/T | [GCKR] | T | 0.416 | 1.04(1.03,1.05) | 1.98X10<sup>-08</sup> |

Chr = chromosome, EAF = Effect allele frequency, OR = Odds ratio, CI = Confidence Interval

The variants, rs75683620, at chr2q12.1 (*TMEM182,* alleles A/G with G being the effect allele, p=5.0 × 10^-09^, **Supplementary Figure 3**), rs536843010 at chr13q33.1 (*NALCN,* alleles A/C with C being the effect allele, p=9.8×10^-09^, **Supplementary Figure 4**), and rs13245319 at chr7p22.3 (*GRIFIN--[]-LFNG*, alleles C/T with T being the effect allele, p = 1.79×10^-08^, **Supplementary Figure 5**) were not associated with functional effects in eQTL or pQTL analysis. These variants are relatively rare in the studied populations, except for rs75683620 (MAF= 0.042) among individuals of African American ancestry. These variants were monomorphic among East and South Asians, meaning that data from these populations did not contribute to the meta-analysis.

We also identified an association at chr2p23.3 (*GCKR*, rs1260326, alleles C/T with T being the effect allele, **Supplementary Figure 6**) that was genome-wide significant in the meta-analysis (p = 1.98×10^-08^) and in the European population (p = 2.0×10^-08^). rs1260326 is a missense variant that is also in high LD with eQTLs for other nearby genes across multiple tissues, including kidney tubules (eQTL gene = *NRBP1*, p=5.3×10^-07^), CD4+ T cells (eQTL gene=*NRBP1*, p=2.8×10^-07^), and liver (eQTL gene=*C2orf16*, p=4.9×10^-23^) (**Supplementary Table 3d**)^15–18^. rs1260326 is also located within 500kb with r^2^ > 0.8 of multiple pQTLs based on data from blood plasma (**Supplementary Table 3b**)^19,20^. As previously reported, rs1260326 is a pleiotropic variant associated with multiple traits ^21^. Similarly to other variants, most of the support for this association also comes from the European population.

### Association with COVID-19 measures

To explore the specificity of these genetic variants with COVID-19+ diarrhea vs COVID-19 severity, we examined the association of the lead variants with COVID-19 susceptibility and COVID-19 severity measures. The variant, rs3504462 (A/G with G as the risk allele), on chr3p21.31 was associated with COVID-19 test positivity (p = 1.1X 10^-05^) and acute COVID-19 infection leading to hospitalization (p = 1.1×10^-56^) or severe respiratory disease (p = 7.1×10^-69^). In contrast, the other variants were not associated with any of these COVID-19 measures (**Supplementary Table 4**). Thus, with the exception of the chr3p21.31 locus, the genetic signals of COVID-19+ diarrhea are likely to drive their effects through mechanisms unrelated to COVID-19 severity.

### PheWAS

To characterize these loci, we performed a phenome-wide association study (PheWAS) across 1,482 phenotypes available in the 23andMe, Inc. database in the population of European ancestry. PheWAS results for variants are presented in **Supplementary Table 5**. Among phenotypes associated with rs10504255, the strongest associations were with high cholesterol (p = 7.53 × 10^-172^) and IBS-D (p = 5.31 × 10^-134^). Other associated phenotypes were cardiometabolic traits such as type 2 diabetes, high blood pressure and statin use (**Supplementary Table 5a**). In addition to the measures of COVID-19 susceptibility and severity, the rs35044562 variant was also associated with autoimmune conditions (Hashimoto’s disease, celiac disease) (**Supplementary Table 5b**). PheWAS for rs1260326 identified associations with lipid profile, allergic conditions, and blood glucose levels (**Supplementary Table 5c**). The main associations for rs1324319 were IBS, kidney stones, and obesity (**Supplementary Table 5d**). No traits were associated with rs75683620 or rs536843010 at the Bonferroni-corrected p-value threshold.

Focusing specifically on gastrointestinal disorders among outcomes constituting the PheWAS analysis, and orienting effect alleles to a higher risk of COVID-19+ diarrhea, four SNPs (rs10504255, rs1260326, rs35044562 and rs13245319) demonstrated associations with GI traits in addition to COVID19+ diarrhea (**Figure 2**), including IBD (both ulcerative colitis and Crohn’s disease), celiac disease, lactose intolerance and IBS. Directionally consistent associations with higher risks of IBS (for rs10504255, rs1260326 and rs13245319) and IBS-D (rs10504255, rs1260326) were identified whereas a directionally opposite association between COVID-19+ diarrhea and IBS-C (rs10504255) was found (**Supplementary Figure 7**).

**Figure 2:**
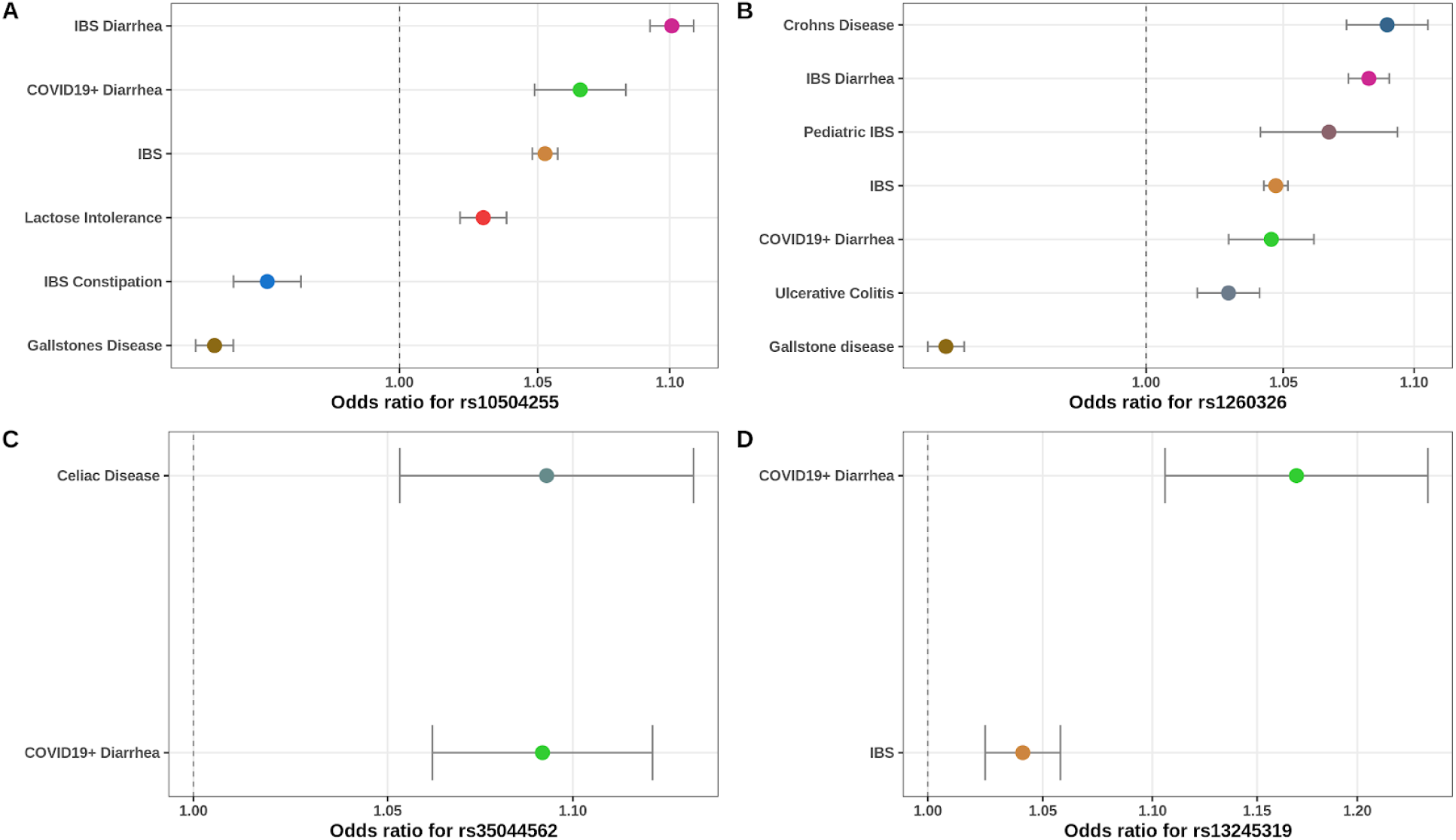
Phenome-wide association of GWAS significant hits of COVID-19+ diarrhea in 23andMe participants of European ancestry. Forest plot represents statistically significant findings from PheWAS analysis of four loci. The results are presented as odds ratio and 95% confidence intervals under additive model for allele of each loci to represent increased odds of having COVID-19+ diarrhea. X-axis shows estimates on log scale. Y-axis shows phenotypes studied. We have included gastrointestinal phenotypes from PheWAS analysis that met the statistically significant threshold of 5.62 × 10^-06^. A complete list of phenotypes that met statistical threshold are included in Supplementary Table 5.

### Mendelian randomization

Given that a common association on PheWAS across three of the six lead variants was an association with IBS and IBS subtypes, and to more fully characterize the translational relevance, we investigated the potential causal relationship between genetic liability to IBS-C and IBS-D and COVID-19+ diarrhea through Mendelian randomization (MR). Using a genetic instrument consisting of 180 SNPs for IBS-D, we identified strong evidence of a potential causal effect of liability to IBS-D and risk of COVID-19+ diarrhea (OR=1.40,95%,CI[1.33-1.47] from random-effects IVW modeling). Steiger filtering removed three SNPs with minimal impact to the IVW MR estimate (OR=1.39,95% CI[1.32-1.46]). These causal estimates persisted or strengthened on robust MR approaches: weighted median provided findings that were largely similar to IVW MR (OR=1.37,95%,CI[1.28-1.47]), with the estimate from MR Egger yielding a stronger predicted causal effect (OR=2.02,95%,CI[1.76-2.32]) (**Figure 3**, **Supplementary Figure 8**). The intercept from MR Egger regression was statistically significantly different from zero [β(SE) = -0.121(0.002); p= 1.74 × 10^-07^], indicating presence of directional pleiotropy (**Supplementary Table 6**).

**Figure 3:**
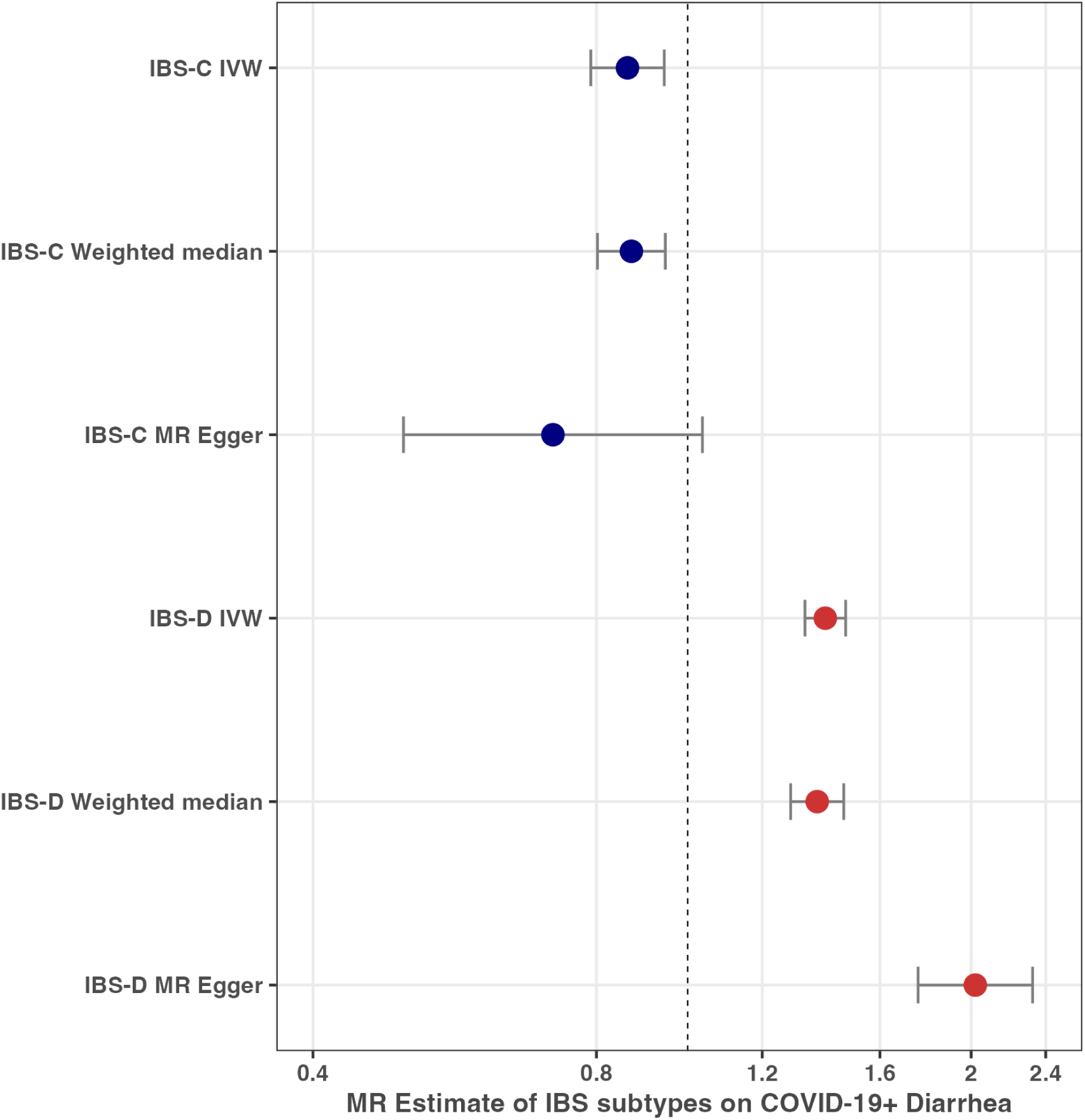
Forest plot representing genetically predicted effects of irritable bowel syndrome subtypes on COVID-19+ diarrhea using Mendelian randomization. The estimates in the plot depict odds ratio and 95% confidence intervals. IBS-C = Irritable bowel syndrome subtype constipation, IBS-D = Irritable bowel syndrome subtype diarrhea, IVW = Inverse variance weighted. The genetic instrument for IBS-C is derived using information from 50 SNPs (meanF statistic = 44.1) and genetic instrument for IBS-D is derived using information from 213 SNPs (meanF statistic = 49.5).

The MR estimates for IBS-C were directionally opposite (0.86, 95%CI[0.79-0.94]; IVW modeling) to those of IBS-D. MR estimates remained unchanged after removing one SNP following Steiger filtering (OR=0.89,95% CI[0.82-0.96]). Robust MR analyses provided consistent findings although the magnitude of the predicted causal estimate was further from the null on MR-Egger; the intercept from MR Egger was β(SE) = 0.008(0.008) with a p-value of 0.31.

Given partial overlap of individuals contributing to GWAS (**Supplementary Figure 9**), we conducted a sensitivity analysis using non-overlapping samples which yielded nearly identical findings (**Supplementary Table 7**).

## DISCUSSION

In this era of widespread documentation of SARS-CoV2 effects on human health, there is an important gap in understanding extrapulmonary symptoms. In this study, we address this gap by utilizing a direct-to-consumer research platform contributing data on 80,289 cases of diarrhea during acute SARS-CoV2 infection. We identified three loci (*CYP7A1*, *LZTFL1–CCR9*, and *GCKR*) that have plausible biological mechanisms of action. Except for *LZTFL1-CCR9*, none of these genetic signals was associated with COVID-19 severity, indicating the role of distinct biological pathways. We further observed a consistent association of top variants at the *CYP7A1*,*GCKR*, and *LFNG* loci with IBS and IBS sub-types. Genetic liability towards IBS-D increased the risk of having COVID-19+ diarrhea, while genetic liability for IBS-C reduced this risk. These results highlight the role of genetic predisposition to preexisting comorbidities in the extrapulmonary manifestations of SARS-CoV2 infection.

The top significant locus in our study, mapped to *CYP7A1*, encodes a member of the cytochrome p450 enzyme family, which has an important role in the bile acid synthesis pathway ^22^. Polymorphisms at this locus are associated with defects in bile acid synthesis, affecting enzymatic activity of cholesterol 7-alpha hydroxylase and resulting in bile acid diarrhea. On eQTL analysis, we observed rs10504255 to be in high LD with a variant (rs8192870) associated with expression of a nearby gene (*SDCBP*, r2=0.93,p=9.9×10-10) in the left ventricular myocardium ^12^. *SDCBP* is also expressed in the intestine, as well as in lung cancers including adenocarcinoma and small cell carcinoma^12^. A recent single-cell RNA sequencing study observed that *SDCBP* likely plays a role in antigen processing and presentation in bronchial epithelial cells from COVID-19 patients, ^13^ with *SDCBP* expression correlating with *HLA-DPB1* expression in normal lung tissues ^13^. *HLA-DPB1* represents a critical immune mechanism as it is expressed on antigen presenting cells that participate in eliciting an immune response to foreign viral peptides. Overall, these findings suggest a possible role of bile acid synthesis pathways and/or the immune system in COVID-19+ diarrhea.

*LZTFL1*, *SLC6A20* and *CCR9* are part of the chemokine receptor gene cluster at chr3p21, previously identified as COVID-19 susceptibility and severity loci ^23–26^. *LZTFL1* at this locus likely regulates viral response pathways by inhibiting the transcription factors that reduce levels of ACE2 and TMPRSS ^14^. *SLC6A20* is extensively expressed in the gastrointestinal tract, where it forms a complex with ACE2 receptors, facilitating viral entry ^24^. *CCR9* is expressed in T-lymphocytes of the small intestine and colon, where it regulates chemokines and eosinophil recruitment ^23,27^. Collectively, these mechanisms may contribute to pathophysiology of diarrheal disease during acute SARS-CoV2 infection.

PheWAS characterization of top GWAS loci identified directionally opposite signals with IBS subtypes. Genetic liability to IBS-D increased, and liability to IBS-C decreased the probability of experiencing diarrhea during the acute phase of SARS-CoV2 infection. These effects could be driven by genetic liability to frequency of bowel motions, modifying the manifestation of diarrhea in the context of an acute infection affecting the GI tract. An alternative explanation might be that individuals at greater liability to IBS-D have altered bowel characteristics that make them more susceptible to GI infection and diarrhea in the context of acute COVID-19 infection. For example, studies have demonstrated that individuals with IBS-D have an altered gut microbiome which could play a role in susceptibility to COVID-19+ diarrhea ^28,29^.

The remaining significantly associated genetic loci do not have direct evidence of involvement in the gastrointestinal tract or SARS-CoV2 infection. *GCKR* is a pleiotropic locus associated with C-reactive protein, fasting plasma glucose levels, and blood cell traits ^21,30^. *GCKR*-mediated effects are driven via regulation of glucokinase enzymes that control the first step of glycolysis. The gene *TMEM182* (rs75683620) at chr2q12.1 has been associated with central obesity and systolic blood pressure in Asian populations ^31,32^. The gene is expressed in heart tissue and regulates its effects via tumor necrosis factor-alpha ^32^. The *NALCN* gene (rs536843010) at chr13q33.1 contributes to physiological processes in the neuromuscular junctions by maintaining resting membrane potential via voltage independent, nonselective cation channels ^33^. It accordingly is involved in control of muscular activity, respiration, and circadian rhythms ^33,34^. The function of the LFNG gene (rs13245319) is regulated via the notch signaling pathway ^35^. The notch pathway maintains the homeostasis of multiple tissues and thus plays a role in cancerous growth, including colorectal adenocarcinoma ^36,37^. These associations require independent validation and additional functional investigations to identify any biological relevance to acute COVID-19+ diarrhea.

Our study has several limitations. European participants contributed 75% of our sample size, so the statistical power to detect genetic associations was driven largely by this group. While the large sample size is an important strength for our study, replication of the GWAS and MR findings by other studies would establish reproducibility and strengthen the findings. Diarrhea cases reported 2 to 3 months after acute infection might be misattributed to COVID-19 as opposed to another source. However, the vast majority (99%) of our cases of COVID-19+ diarrhea occurred at the same time as the diagnosis of COVID-19. Furthermore, individuals with genetic liability to IBS-D may have misattributed diarrhea at the time of acute COVID-19 to the infection as opposed to their underlying IBS-D. Refuting this hypothesis is our observation that individuals with IBS-C had a lower relative odds of experiencing COVID-19+ diarrhea, which collectively may be indicative of an acute infection operating on top of a background genetic liability to IBS-D or IBS-C, with liability to either IBS-D or IBS-C influencing the probability of experiencing COVID-19+ diarrhea in a potentially etiological way. Finally, the MR-Egger intercept for IBS-D indicated evidence of directional pleiotropy. However, the pleiotropy-corrected point estimate from MR-Egger was larger (OR=2.02) than that derived from IVW (OR=1.40), making the latter a conservative estimate.

In conclusion, the first GWAS of COVID-19+ diarrhea in an ancestrally diverse, large-scale population-based cohort identifies biologically relevant signals associated with bile acid synthesis and immune function, including antigen presenting and cytokine signaling. By virtue of limiting our study to COVID-19+ individuals, we infer and substantiate empirically that all but one of the identified loci are specific to COVID-19+ diarrhea rather than markers of susceptibility to SARS-CoV2 infection. Finally, we provide causal evidence in support of the etiological role of liability to chronic intestinal disorders and gastrointestinal symptoms during acute infection.

## METHODS

### Study Population

Participants older than 18 years old were recruited for 23andMe COVID-19 study from April 2020 using email-based surveys. The surveys were distributed to 6.7 million individuals who provided informed consent and volunteered to participate in the research online, under a protocol approved by the external AAHRPP-accredited IRB, Ethical & Independent (E&I) Review Services. As of 2022, E&I Review Services is part of Salus IRB. Recruitment was initially geo-targeted to capture cases as the outbreak spread across the U.S and was continued later on to recruit additional participants. We included only those participants who had responded to COVID-19 survey and provided symptom information by August, 2023 for this study. Details on diagnosis, testing, and symptoms of COVID-19, as well as markers of severity and relevant comorbid conditions were collected via one baseline and three follow-up surveys administered three months apart. Full details of the data collection procedures for this study have been described previously^38^.

### Phenotype

Using the survey information, we defined the diarrhea outcome among individuals who self-reported testing positive for COVID-19. Participants were asked the question whether they experienced any of the following symptoms to which they could select as many as needed from the following list of responses: ‘muscle or body aches/ fatigue/ dry cough/ sore throat/ coughing up of sputum or phlegm (productive cough)/ loss of smell or taste/ chills/ difficulty breathing or shortness of breath/ pressure or tightness in upper chest/ diarrhea/ nausea or vomiting/ sneezing/ loss of appetite/ runny nose/ headache/ intensely red or watery eyes’. Participants who reported experiencing diarrhea symptoms at baseline or follow-up surveys were identified as cases and those who did not experience diarrhea were controls. The analytical dataset included unrelated individuals with non-missing information on COVID-19+ diarrhea and covariates included in GWAS analyses (N=239,866).

### Genotyping

DNA extraction and genotyping were performed on saliva samples by Clinical Laboratory Improvement Amendments-certified and College of American Pathologists-accredited clinical laboratories of Laboratory Corporation of America. Samples were genotyped across the five genotyping platforms and imputed using three combined independent reference panels: the publicly available Human Reference Consortium (HRC), and UK BioBank (UKBB) 200K Whole Exome Sequencing (WES) reference panels and the 23andMe reference panel, which was built by 23andMe using internal and external cohorts. Each genotyping platform was imputed and phased separately. The final genotyped variants included 1,469,237 variants and the final imputation panel included a total of 99,675,338 variants (90,582,19 SNPs and 9,093,144 indels). The variant quality control statistics were computed independently with each phasing panel and genotyping platform. We removed variants with low imputation quality (r2 < 0.5 averaged across batches or a minimum r2 < 0.3) or evidence of differences in effects across batches. For genotyped variants, we removed variants only present on our V1 or V2 arrays (due to small sample size) that failed a Mendelian transmission test in trios (P < 10^−20^), failed a Hardy–Weinberg test in individuals of European ancestry (P < 10^−20^), failed a batch effect test (ANOVA P < 10^−50^) or had a call rate <90%. For imputed variants in HRC panel, following filters were used: singletons were excluded, multi-allelics were split into bi-allelic variants (with bcftools), variants with >20% missingness were removed, variants with minor allele count == 0 were removed, and variants with inbreeding coefficient < -0.3 (high heterozygosity) were removed.

### Ancestry Classification

Ancestries in the 23andMe database are determined using a classifier algorithm based on analysis of local ancestry ^39^. Phased genotyped data were first partitioned into windows of about 300 SNPs and a support vector machine (SVM) approach was applied within each window to classify individual haplotypes into one of 45 worldwide reference populations. The SVM classifications are then fed into a hidden Markov model (HMM) that accounts for switch errors and incorrect assignments, and gives probabilities for each reference population in each window. Finally, we used simulated admixed individuals to recalibrate the HMM probabilities so that the reported assignments are consistent with the simulated admixture proportions. We aggregated the probabilities of the 45 reference populations into six main ancestries (European, African-American, Latinx, East Asian, South Asian, Middle Eastern) using a predetermined threshold ^39^. African Americans and Latinx were admixed with broadly varying contributions from Europe, Africa and the Americas. No single threshold of genome-wide ancestry could effectively discriminate between African Americans and Latinx. However, the distributions of the length of segments of European, African and American ancestry are very different between African Americans and Latinx, because of distinct admixture timing between the three ancestral populations in the two ethnic groups. Therefore, we trained a logistic classifier that took the participant’s length histogram of segments of African, European and American ancestry, and predicted whether the customer is likely African American or Latinx.

### GWAS Analysis

To obtain unrelated participants for our GWAS analyses, individuals were included such that no two individuals shared more than 700 cM of DNA identical by descent. We excluded approximately 1.80% of the sample to obtain such a set of unrelated individuals. If a case and control were identified as having at least 700cM of DNA IBD, we retained the case from the sample. We conducted association analysis using logistic regression, assuming additive allelic effects and adjusting for age, age-squared, sex, sex-age interaction, genotyping platform variables and ten principal components to account for residual population structure. We combined the GWAS summary statistics from both genotyped and imputed data. When choosing between imputed and genotyped GWAS results, we favored the imputed result, unless the imputed variant was unavailable or failed quality control. The summary statistics were adjusted for inflation using genomic control when the inflation factor was estimated to be greater than one (λ = 1.029, 1.037, 1.024, 1.042 and 1.071 within the European, Latinx, African American, East Asian and South Asian ancestry GWAS, respectively). We defined the region boundaries by identifying all SNPs with P < 10^−5^ within the vicinity of a genome-wide significance association and then grouping these regions into intervals so that no two regions were separated by less than 250 kb. We considered the SNP with the smallest P value within each interval to be the **index variant.** Within each region, we calculated a credible set of variants using the method of Maller et al 2021 ^40^.

We conducted the GWAS analysis separately in five population cohorts (European, Latinx, African-Americans, East Asian, and South Asian ancestry). We then meta-analyzed the GWAS summary statistics of these populations using an inverse variance fixed effect model. For this approach, we included variants that had at least 1% minor allele frequency in the pooled sample and minor allele count > 30 within each subpopulation. The resulting meta-analyses were also adjusted for inflation (λ = 1.001) using genomic control.

### PheWAS Analysis

We conducted phenome-wide associations (PheWAS) on the index variants from loci that were statistically significant on multi-ancestry analysis. The PheWAS analysis was limited to data from participants of European ancestry. We used data on 1,482 phenotypes that were available in the 23andMe research database. Data on these phenotypes were collected using online survey-based questionnaires completed by the participants at the time of recruitment in 23andMe genetic database. PheWAS analysis was performed, adjusting the association between each phenotype and variant of interest for age, sex, and the first five principal components of ancestry. We reported the associations that met the statistical threshold of significance after correcting for multiple testing (p < 0.05 / (1,482*6)= 5.62 × 10^-06^).

### Mendelian Randomization

Because PheWAS highlighted a shared association with irritable bowel syndrome (IBS) and IBS subtypes, we explored whether there was a potential causal role of genetic liability to IBS subtypes and COVID-19+ diarrhea by conducting two-sample Mendelian randomization (MR) using the *twosampleMR* R package ^41^. The initial MR analysis included overlapping samples of IBS subtypes and COVID-19+ diarrhea in the European population. We focused on the IBS subtypes of diarrhea (IBS-D) and constipation (IBS-C). We first obtained separate genetic instruments for IBS-C and IBS-D from genome-wide significant SNPs for the relevant endpoint that had a minor allele frequency of more than 0.01. We then identified variants with the smallest p-value (index variants) by defining the regions of significant association based on genome-wide significant SNPs as described above. These index variants were included in the genetic instrument of the relevant phenotype (IBS-C or IBS-D). The genetic instrument for IBS-D included 180 SNPs (mean F-statistic per SNP=50.2) and for IBS-C included 52 SNPs (mean F-statistic per SNP=43.6). Using the summary statistics of SNPs with IBS-D and IBS-C adjusted for age, sex, and principal components of ancestral structure in the European population, we then fitted random effects inverse variance weighted (IVW) models and obtained MR estimates of IBS subtypes on COVID-19+ diarrhea. To test the veracity of the findings, we used Steiger filtering and robust MR approaches including weighted median, and MR-Egger method ^41–43^. As a sensitivity analysis, we repeated MR analysis between IBS subtypes and COVID-19+ diarrhea after excluding the overlapping samples between them.

### Functional Annotation of GWAS Index Variants

To perform variant-to-gene mapping, hypotheses of functionally relevant genes are generated by annotating the strongest associations (index variants) with nearby functional variants. The mapping is computed by searching functional variants within 500 kb of the index variant with a filter of linkage disequilibrium r2 > 0.8. Functional variants for mapping include coding variants (annotated by the Ensembl Variant Effect Predictor (VEP) v109 ^44^), eQTLs, and pQTLs. The eQTL annotation resources consist of a comprehensive collection of standardized variants impacting gene expression in various tissues obtained from publicly available datasets ^12,15–18,45–48^ and datasets processed by the 23andMe eQTL pipeline. The pQTL annotation resources similarly include a collection of curated protein QTLs from relevant public datasets ^19,20^. *eQTL discovery*. eQTL calling was performed with one of two versions of 23andMe pipelines, depending on the dataset in question (**Supplementary File**). The first pipeline used FastQTL ^49^ in permutation mode, restricting all tests to variants within a window defined to be 1Mbp up- or downstream of a given gene’s transcription start site (TSS) (**Supplementary File Table 1**). Variants tested were single nucleotide polymorphisms with an in-sample MAF ≥ 1%, to avoid errors in detection or mapping of larger genetic variants in cross-ancestry comparisons, and models were adjusted for age (if available), sex, probabilistic estimation of expression residuals (PEER) factors ^50^, genetic PCs, and per-dataset covariates. For each gene, the index variant was identified by the minimal permutation p-value. eQTLs were called then on lead variants if they passed a 5% FDR filter using Storey’s q-value ^51^ methodology. Conditional eQTLs were identified via FastQTL’s permutation mode, by using each eQTL as an additional covariate in the model for a given gene. The lead conditional eQTL for all genes were again FDR controlled at 5%, and a maximum of 10 conditional steps were run. Finally, for a set of conditional eQTLs for a given gene, a joint model was fit, and the final eQTL callset consisted of those eQTLs whose joint model test passed a 5% FDR filter. eQTLs were only called for genes classified as one of ‘protein_coding’, ‘miRNA’, ‘IG_C_gene’, ‘IG_D_gene’, ‘IG_J_gene’, ‘IG_V_gene’,’TR_C_gene’, ‘TR_D_gene’, ‘TR_J_gene’, ‘TR_V_gene’ as defined in GENCODE^52^.

The second version of the 23andMe pipeline uses strand-aware RNA-seq quantification, and the eQTLs were called using SusieR package ^53^ instead of FastQTL, with expression PCs (selected with the elbow method) replacing PEER factors in the modeling, and using the GENCODE v43 gene model (**Supplementary File Table 2**). The pipeline natively generated credible sets with a set probability to contain a SNP tagging the causal variant.

## DATA AVAILABILITY

The full set of GWAS summary statistics can be made available to qualified investigators upon request and signing agreement with 23andMe to protect participant confidentiality. The information can be accessed at https://research.23andme.com/covid19-dataset-access/

## ACKNOWLEDGEMENTS

We thank the 23andMe research participants and employees who made this study possible. The following members of the 23andMe Research Team contributed to this study:

Stella Aslibekyan, Adam Auton, Elizabeth Babalola, Robert K. Bell, Jessica Bielenberg, Jonathan Bowes, Katarzyna Bryc, Ninad S. Chaudhary, Daniella Coker, Sayantan Das, Emily DelloRusso, Sarah L. Elson, Nicholas Eriksson, Teresa Filshtein, Pierre Fontanillas, Will Freyman, Zach Fuller, Chris German, Julie M. Granka, Karl Heilbron, Alejandro Hernandez, Barry Hicks, David A. Hinds, Ethan M. Jewett, Yunxuan Jiang, Katelyn Kukar, Alan Kwong, Yanyu Liang, Keng-Han Lin, Bianca A. Llamas, Matthew H. McIntyre, Steven J. Micheletti, Meghan E. Moreno, Priyanka Nandakumar, Dominique T. Nguyen, Jared O’Connell, Aaron A. Petrakovitz, G. David Poznik, Alexandra Reynoso, Shubham Saini, Morgan Schumacher, Leah Selcer, Anjali J. Shastri, Janie F. Shelton, Jingchunzi Shi, Suyash Shringarpure, Qiaojuan Jane Su, Susana A. Tat, Vinh Tran, Joyce Y. Tung, Xin Wang, Wei Wang, Catherine H. Weldon, Peter Wilton, Corinna D. Wong.

## AUTHOR INFORMATION

### Affiliations

**23andMe, Inc:**

Ninad S. Chaudhary, Catherine H. Weldon, Priyanka Nandakumar, 23andMe Research Team, Michael V. Holmes, Stella Aslibekyan

**Bristol Myers Squibb, Inc:**

Janie F. Shelton

## Author Contributions

The 23andMe COVID-19 Team developed the recruitment and participant engagement strategy and acquired and processed the data. N.S.C analyzed the data. N.S.C., P.A, C.H.W, S.A, and M.V.H interpreted the data. N.S.C., S.A. and M.V.H. wrote the manuscript. All authors participated in the preparation of the manuscript by reading and commenting on the drafts before submission.

## ETHICS DECLARATIONS

## Competing Interests

C.H.W, P.N, S,A, and M.V.H are current employees of 23andMe and hold stock or stock options in 23andMe. J.S is a current employee of Bristol Myers Squibb. N.S.C works as a postdoctoral fellow on the 23andMe Genetic Epidemiology Team.

## Notes

### Funding Statement

This study did not receive any funding

### Author Declarations

The surveys were distributed to individuals who provided informed consent and volunteered to participate in the research online, under a protocol approved by the external AAHRPP-accredited IRB, Ethical & Independent (E&I) Review Services. As of 2022, E&I Review Services is part of Salus IRB

### Summary of Updates

MR Egger estimates changed in the result section Figure 3 and Supplementary Figure 8 revised for updated MR Egger estimates

## REFERENCES

1. Mao, R. et al. Manifestations and prognosis of gastrointestinal and liver involvement in patients with COVID-19: a systematic review and meta-analysis. Lancet Gastroenterol. Hepatol. 5, 667–678 (2020).

2. Parasa, S. et al. Prevalence of Gastrointestinal Symptoms and Fecal Viral Shedding in Patients With Coronavirus Disease 2019: A Systematic Review and Meta-analysis. *JAMA Netw*. Open 3, e2011335 (2020).

3. Sultan, S. et al. AGA Institute Rapid Review of the Gastrointestinal and Liver Manifestations of COVID-19, Meta-Analysis of International Data, and Recommendations for the Consultative Management of Patients with COVID-19. Gastroenterology 159, 320–334.e27 (2020).

4. Nardo, A. D. et al. Pathophysiological mechanisms of liver injury in COVID-19. Liver Int. Off. J. Int. Assoc. Study Liver 41, 20–32 (2021).

5. Lin, L. et al. Gastrointestinal symptoms of 95 cases with SARS-CoV-2 infection. Gut 69, 997–1001 (2020).

6. Xu, E., Xie, Y. & Al-Aly, Z. Long-term gastrointestinal outcomes of COVID-19. Nat. Commun. 14, 983 (2023).

7. Natarajan, A. et al. Gastrointestinal symptoms and fecal shedding of SARS-CoV-2 RNA suggest prolonged gastrointestinal infection. Med N. Y. N 3, 371–387.e9 (2022).

8. Hoffmann, M. et al. SARS-CoV-2 Cell Entry Depends on ACE2 and TMPRSS2 and Is Blocked by a Clinically Proven Protease Inhibitor. Cell 181, 271–280.e8 (2020).

9. Hamming, I. et al. Tissue distribution of ACE2 protein, the functional receptor for SARS coronavirus. A first step in understanding SARS pathogenesis. J. Pathol. 203, 631–637 (2004).

10. Zollner, A. et al. Postacute COVID-19 is Characterized by Gut Viral Antigen Persistence in Inflammatory Bowel Diseases. Gastroenterology 163, 495–506.e8 (2022).

11. Stein, S. R. et al. SARS-CoV-2 infection and persistence in the human body and brain at autopsy. Nature 612, 758–763 (2022).

12. GTEx Consortium. The GTEx Consortium atlas of genetic regulatory effects across human tissues. Science 369, 1318–1330 (2020).

13. Ma, D. et al. Single-cell RNA sequencing identify SDCBP in ACE2-positive bronchial epithelial cells negatively correlates with COVID-19 severity. J. Cell. Mol. Med. 25, 7001–7012 (2021).

14. Downes, D. J. et al. Identification of LZTFL1 as a candidate effector gene at a COVID-19 risk locus. Nat. Genet. 53, 1606–1615 (2021).

15. Qiu, C. et al. Renal compartment-specific genetic variation analyses identify new pathways in chronic kidney disease. Nat. Med. 24, 1721–1731 (2018).

16. Kettunen, J. et al. Genome-wide association study identifies multiple loci influencing human serum metabolite levels. Nat. Genet. 44, 269–276 (2012).

17. Franzén, O. et al. Cardiometabolic risk loci share downstream cis- and trans-gene regulation across tissues and diseases. Science 353, 827–830 (2016).

18. Raj, T. et al. Polarization of the effects of autoimmune and neurodegenerative risk alleles in leukocytes. Science 344, 519–523 (2014).

19. Zhang, J. et al. Plasma proteome analyses in individuals of European and African ancestry identify cis-pQTLs and models for proteome-wide association studies. Nat. Genet. 54, 593–602 (2022).

20. Ferkingstad, E. et al. Large-scale integration of the plasma proteome with genetics and disease. Nat. Genet. 53, 1712–1721 (2021).

21. Orho-Melander, M. et al. Common missense variant in the glucokinase regulatory protein gene is associated with increased plasma triglyceride and C-reactive protein but lower fasting glucose concentrations. Diabetes 57, 3112–3121 (2008).

22. Chiang, J. Y. L. & Ferrell, J. M. Up to date on cholesterol 7 alpha-hydroxylase (CYP7A1) in bile acid synthesis. Liver Res. 4, 47–63 (2020).

23. Pathak, M. & Lal, G. The Regulatory Function of CCR9+ Dendritic Cells in Inflammation and Autoimmunity. Front. Immunol. 11, 536326 (2020).

24. Severe Covid-19 GWAS Group et al. Genomewide Association Study of Severe Covid-19 with Respiratory Failure. N. Engl. J. Med. 383, 1522–1534 (2020).

25. Shelton, J. F. et al. Trans-ancestry analysis reveals genetic and nongenetic associations with COVID-19 susceptibility and severity. Nat. Genet. 53, 801–808 (2021).

26. COVID-19 Host Genetics Initiative. Mapping the human genetic architecture of COVID-19. Nature 600, 472–477 (2021).

27. Uehara, S., Grinberg, A., Farber, J. M. & Love, P. E. A role for CCR9 in T lymphocyte development and migration. J. Immunol. Baltim. Md 1950 168, 2811–2819 (2002).

28. Menees, S. & Chey, W. The gut microbiome and irritable bowel syndrome. F1000Research 7, F1000 Faculty Rev-1029 (2018).

29. Wang, B. et al. Alterations in microbiota of patients with COVID-19: potential mechanisms and therapeutic interventions. Signal Transduct. Target. Ther. 7, 143 (2022).

30. Dupuis, J. et al. New genetic loci implicated in fasting glucose homeostasis and their impact on type 2 diabetes risk. Nat. Genet. 42, 105–116 (2010).

31. Kim, Y. K. et al. Identification of a genetic variant at 2q12.1 associated with blood pressure in East Asians by genome-wide scan including gene-environment interactions. BMC Med. Genet. 15, 65 (2014).

32. Ma, M., Lee, J. H. & Kim, M. Identification of a TMEM182 rs141764639 polymorphism associated with central obesity by regulating tumor necrosis factor-α in a Korean population. J. Diabetes Complications 34, 107732 (2020).

33. Cochet-Bissuel, M., Lory, P. & Monteil, A. The sodium leak channel, NALCN, in health and disease. Front. Cell. Neurosci. 8, 132 (2014).

34. Bourque, D. K. et al. Periodic breathing in patients with NALCN mutations. J. Hum. Genet. 63, 1093–1096 (2018).

35. Moloney, D. J. et al. Fringe is a glycosyltransferase that modifies Notch. Nature 406, 369–375 (2000).

36. Zhou, B. et al. Notch signaling pathway: architecture, disease, and therapeutics. Signal Transduct. Target. Ther. 7, 95 (2022).

37. Del Castillo Velasco-Herrera, M., et al. Comparative genomics reveals that loss of lunatic fringe (LFNG) promotes melanoma metastasis. Mol. Oncol. 12, 239–255 (2018).

38. Shelton, J. F. et al. The UGT2A1/UGT2A2 locus is associated with COVID-19-related loss of smell or taste. Nat. Genet. 54, 121–124 (2022).

39. Durand, E. Y., Do, C. B., Mountain, J. L. & Macpherson, J. M. Ancestry Composition: A Novel, Efficient Pipeline for Ancestry Deconvolution. http://biorxiv.org/lookup/doi/10.1101/010512 (2014) doi:10.1101/010512.

40. Wellcome Trust Case Control Consortium et al. Bayesian refinement of association signals for 14 loci in 3 common diseases. Nat. Genet. 44, 1294–1301 (2012).

41. Bowden, J., Davey Smith, G., Haycock, P. C. & Burgess, S. Consistent Estimation in Mendelian Randomization with Some Invalid Instruments Using a Weighted Median Estimator. Genet. Epidemiol. 40, 304–314 (2016).

42. Bowden, J., Davey Smith, G. & Burgess, S. Mendelian randomization with invalid instruments: effect estimation and bias detection through Egger regression. Int. J. Epidemiol. 44, 512–525 (2015).

43. Hemani, G., Tilling, K. & Davey Smith, G. Orienting the causal relationship between imprecisely measured traits using GWAS summary data. PLoS Genet. 13, e1007081 (2017).

44. McLaren, W. et al. The Ensembl Variant Effect Predictor. Genome Biol. 17, 122 (2016).

45. Kerimov, N. et al. A compendium of uniformly processed human gene expression and splicing quantitative trait loci. Nat. Genet. 53, 1290–1299 (2021).

46. Lappalainen, T. et al. Transcriptome and genome sequencing uncovers functional variation in humans. Nature 501, 506–511 (2013).

47. Koolpe, G. A. et al. Opioid agonists and antagonists. 6-Desoxy-6-substituted lactone, epoxide, and glycidate ester derivatives of naltrexone and oxymorphone. J. Med. Chem. 28, 949–957 (1985).

48. Craig, D. W. et al. RNA sequencing of whole blood reveals early alterations in immune cells and gene expression in Parkinson’s disease. Nat. Aging 1, 734–747 (2021).

49. Ongen, H., Buil, A., Brown, A. A., Dermitzakis, E. T. & Delaneau, O. Fast and efficient QTL mapper for thousands of molecular phenotypes. Bioinformatics 32, 1479–1485 (2016).

50. Stegle, O., Parts, L., Piipari, M., Winn, J. & Durbin, R. Using probabilistic estimation of expression residuals (PEER) to obtain increased power and interpretability of gene expression analyses. Nat. Protoc. 7, 500–507 (2012).

51. Storey, J. D. A Direct Approach to False Discovery Rates. J. R. Stat. Soc. Ser. B Stat. Methodol. 64, 479–498 (2002).

52. Harrow, J. et al. GENCODE: the reference human genome annotation for The ENCODE Project. Genome Res. 22, 1760–1774 (2012).

53. Wang, G., Sarkar, A., Carbonetto, P. & Stephens, M. A Simple New Approach to Variable Selection in Regression, with Application to Genetic Fine Mapping. J. R. Stat. Soc. Ser. B Stat. Methodol. 82, 1273–1300 (2020).

