## Supplementary Tables and Figures for "Multi-ancestry GWAS of diarrhea during acute SARS-CoV2 infection identifies multiple novel loci and contrasting etiological roles of irritable bowel syndrome subtypes"

| Supplementary Table 1: Association of GWAS significant variants with COVID19+ diarrhea using data from survey collected at survey time points | | | | | | | | | | | | | | |
|---|---|---|---|---|---|---|---|---|---|---|---|---|---|---|
| SNP | | | Baseline (Cases N=79,361, Control = 159,577) | | | 1 Month follow-up (Cases N =313, Control=159,577) | | | 2 Month follow-up (Cases N =346, Control=159,577) | | | 3 Month follow-up (Cases N =269, Control=159,577) | | |
| | Effect allele | EAF | Effect | Stderr | P value | Effect | Stderr | P value | Effect | Stderr | P value | Effect | Stderr | P value |
| rs10504255 | G | 0.344 | -0.06 | 0.01 | 3.99E-18 | -0.16 | 0.09 | 0.0648 | 0.13 | 0.08 | 0.1066 | -0.01 | 0.09 | 0.9524 |
| rs35044562 | G | 0.078 | 0.08 | 0.01 | 9.15E-12 | 0.00 | 0.16 | 0.9984 | -0.12 | 0.16 | 0.4464 | -0.30 | 0.19 | 0.1189 |
| rs1260326 | T | 0.416 | 0.04 | 0.01 | 1.48E-08 | 0.09 | 0.08 | 0.2584 | 0.03 | 0.08 | 0.7378 | 0.12 | 0.09 | 0.1587 |
| rs75683620 | G | 0.999 | -0.23 | 0.06 | 7.55E-05 | -0.53 | 0.64 | 0.4069 | 0.56 | 1.07 | 0.5986 | 35.16 | 56.17 | 0.5313 |
| rs536843010 | C | 0.991 | -0.22 | 0.03 | 3.04E-10 | -0.43 | 0.38 | 0.261 | -0.03 | 0.45 | 0.9553 | 0.31 | 0.59 | 0.6037 |
| rs13245319 | T | 0.021 | 0.15 | 0.02 | 4.34E-10 | -0.32 | 0.38 | 0.4005 | -0.13 | 0.33 | 0.6914 | 0.18 | 0.33 | 0.5797 |
| **Stderr**= standard error; **Effect** is represented on log odds; **EAF** = effect allele frequency | | | | | | | | | | | | | | |

| Supplementary Table 2a : Association of GWAS meta-analysis hits with COVID-19+ diarrhea in European Population | | | | | |
|---|---|---|---|---|---|
| **SNP** | **Alleles** | **Effect allele** | **EAF** | **OR(95% CI)** | **P value** |
| rs10504255 | A/G | G | 0.344 | 0.94 (0.92, 0.95) | 1.08E-14 |
| rs35044562 | A/G | G | 0.078 | 1.09 (1.06, 1.12) | 1.44E-09 |
| rs75683620 | A/G | G | 0.999 | 0.50 (0.32, 0.81) | 4.42E-03 |
| rs536843010 | A/C | C | 0.991 | 0.81 (0.74, 0.87) | 7.88E-08 |
| rs13245319 | C/T | T | 0.021 | 1.17 (1.10, 1.24) | 9.31E-08 |
| rs1260326 | C/T | T | 0.416 | 1.05 (1.03,1.06) | 2.03E-08 |

**EAF =** effect allele frequency**; OR** = odds ratio; **CI** = Confidence intervals

**Supplementary Table 2b: Association of GWAS meta-analysis hits with COVID-19+ diarrhea in African American Population**

| SNP | Alleles | Effect allele | EAF | OR (95% CI) | P value |
|---|---|---|---|---|---|
| rs10504255 | A/G | G | 0.113 | 0.90 (0.81 , 1.00) | 5.33E-02 |
| rs35044562 | A/G | G | 0.022 | 1.01 (0.81, 1.25) | 9.42E-01 |
| rs75683620 | A/G | G | 0.958 | 0.69 (0.59, 0.82) | 1.30E-05 |
| rs536843010 | A/C | C | 0.997 | 0.93 (0.53, 1.62) | 8.03E-01 |
| rs13245319 | C/T | T | 0.006 | 1.20 (0.80, 1.81) | 3.80E-01 |
| rs1260326 | C/T | T | 0.179 | 1.05 (0.97, 1.15) | 2.46E-01 |

**EAF =** effect allele frequency, **OR** = odds ratio, **CI** = Confidence intervals

| **Supplementary Table 2c : Association of GWAS meta-analysis hits with COVID-19+ diarrhea in Latinx Population** | | | | | |
|---|---|---|---|---|---|
| **SNP** | **Alleles** | **Effect allele** | **EAF** | **OR (95% CI)** | **P value** |
| rs35044562 | A/G | G | 0.057 | 1.13 (1.06, 1.21) | 2.86E-04 |
| rs75683620 | A/G | G | 0.997 | 0.67 (0.51, 0.88) | 4.57E-03 |
| rs536843010 | A/C | C | 0.995 | 0.80 (0.64, 1.00) | 5.43E-02 |
| rs13245319 | C/T | T | 0.015 | 1.12 (0.97, 1.28) | 1.16E-01 |
| rs1260326 | C/T | T | 0.369 | 1.02 (0.99, 1.06) | 1.64E-01 |
| **EAF** = effect allele frequency, **OR** = odds ratio, **CI** = Confidence intervals | | | | | |

**Supplementary Table 2d : Association of GWAS meta-analysis hits with COVID-19+ diarrhea in East Asian Population**

| SNP | Alleles | Effect allele | EAF | OR (95% CI) | P value |
|---|---|---|---|---|---|
| rs10504255 | A/G | G | 0.215 | 0.89 (0.78, 1.01) | 6.54E-02 |
| rs35044562 | A/G | G | 0.011 | 1.57 (1.07, 2.30) | 2.24E-02 |
| rs1260326 | C/T | T | 0.489 | 1.02 (0.92, 1.14) | 6.51E-01 |

**EAF** = effect allele frequency, **OR** = odds ratio, **CI** = Confidence intervals

**Supplementary Table 2e : Association of GWAS meta-analysis hits with COVID-19+ diarrhea in South Asian Population**

| SNP | Alleles | Effect allele | EAF | OR (95% CI) | P value |
|---|---|---|---|---|---|
| rs10504255 | A/G | G | 0.340 | 0.84 (0.69, 1.03) | 8.90E-02 |
| rs35044562 | A/G | G | 0.275 | 1.16 (0.94, 1.42) | 1.78E-01 |
| rs1260326 | C/T | T | 0.228 | 0.76 (0.62, 0.95) | 1.36E-02 |

**EAF** = effect allele frequency, **OR** = odds ratio, **CI** = Confidence intervals

**Supplementary Table 3a: Coding signals identified on variant to gene effects for GWAS hits in European ancestry population**

| GWAS SNP | P value | Gene | Consequence | r square | Coding SNP | Population |
|---|---|---|---|---|---|---|
| rs1260326 | 2.03E-08 | GCKR | missense_variant&splice_region_variant | 1 | rs1260326 | european |

**r square** = LD correlation coefficient

**Supplementary Table 3b: Expression quantitative trait locus signals identified on variant to gene effects for GWAS hits in European and Latino ancestry population**

| GWAS SNP | eQTL SNP | eqtl Gene | eqtl tissue type | eqtl P value | r square of GWAS and eQTL SNP | Population |
|---|---|---|---|---|---|---|
| rs1260326 | rs4665972 | NRBP1 | renal_tubule | 5.27E-07 | 0.93 | european |
| rs1260326 | rs4665972 | NRBP1 | naive_CD4_T_cell | 2.77E-07 | 0.93 | european |
| rs1260326 | rs1260326 | C2orf16 | liver | 4.93E-23 | 1.00 | european |
| rs35896106 | rs17712877:C | SLC6A20 | breast_epithelium | 1.79E-07 | 0.92 | european |
| rs10504255 | rs9297994 | CYP7A1 | thyroid_gland | 8.06E-09 | 0.97 | european |
| rs10504255 | rs8192870 | SDCBP | left_ventricle_myocardium | 9.90E-10 | 0.94 | european |
| rs12335362 | rs12348713 | SMC2 | B_cell | 6.29E-10 | 0.85 | latinx |
| rs12335362 | rs12335362 | SMC2 | CD14_positive_monocyte_activated_24h_IFNg | 2.58E-19 | 1.00 | latinx |
| rs12335362 | rs12335362 | SMC2 | NK_cell | 1.43E-15 | 1.00 | latinx |

**eqtl** = expression quantitative trait locus; **r square** = LD correlation coefficient

| Supplementary Table 3c: Protein quantitative trait locus signals identified on variant to gene effects for GWAS hits in European ancestry population | | | | | | |
|---|---|---|---|---|---|---|
| **GWAS SNP** | **pqtl SNP** | **pqtl Gene** | **pqtl tissue type** | **pqtl P value** | **rsquare between GWAS and pQTL SNP** | **population** |
| rs1260326 | rs1260326 | GCKR | blood_plasma | 1.35E-31 | 1 | european |
| rs1260326 | rs1260326 | NRXN3 | blood_plasma | 1.20E-15 | 1 | european |
| rs1260326 | rs1260326 | KNG1 | blood_plasma | 8.91E-10 | 1 | european |
| rs1260326 | rs1260326 | INHBA | blood_plasma | 4.47E-33 | 1 | european |
| rs1260326 | rs1260326 | NTM | blood_plasma | 1.95E-29 | 1 | european |
| rs1260326 | rs1260326 | F9 | blood_plasma | 2.09E-26 | 1 | european |
| **pqtl** = protein quantitative trait locus; **r square** = LD correlation coefficient | | | | | | |

| Supplementary Table 3d: Expression quantitative trait locus signals identified on variant to gene effects for GWAS hits from meta-analysis findings | | | | | | |
|---|---|---|---|---|---|---|
| **GWAS SNP** | **eQTL SNP** | **eqtl Gene** | **eqtl tissue type** | **eqtl P value** | **r square between GWAS and eQTL SNP** | **population** |
| rs1260326 | rs4665972 | NRBP1 | renal_tubule | 5.27E-07 | 0.93 | european |
| rs1260326 | rs4665972 | NRBP1 | naive_CD4_T_cell | 2.77E-07 | 0.93 | european |
| rs1260326 | rs1260326 | C2orf16 | liver | 4.93E-23 | 1.00 | european |
| rs35044562 | rs17712877:C | SLC6A20 | breast_epithelium | 1.79E-07 | 0.93 | european |
| rs35044562 | rs73064431 | SLC6A20 | esophagus_muscularis_mucosa | 1.47E-08 | 0.89 | european |
| rs10504255 | rs9297994 | CYP7A1 | thyroid_gland | 8.06E-09 | 0.97 | european |
| rs10504255 | rs8192870 | SDCBP | left_ventricle_myocardium | 9.90E-10 | 0.94 | european |
| rs910888 | rs4413223 | KIF3B | renal_tubule | 1.41E-08 | 0.94 | european |
| rs910888 | rs6119231 | KIF3B | tibial_artery | 3.51E-09 | 0.94 | european |
| rs910888 | rs3903368 | KIF3B | lymphoblast | 4.55E-07 | 0.95 | european |
| rs910888 | rs1742980 | KIF3B | esophagus_muscularis_mucosa | 5.03E-08 | 0.96 | european |
| rs1260326 | rs4665972 | NRBP1 | renal_tubule | 5.27E-07 | 0.89 | latinx |
| rs1260326 | rs4665972 | NRBP1 | naive_CD4_T_cell | 2.77E-07 | 0.89 | latinx |
| rs1260326 | rs1260326 | C2orf16 | liver | 4.93E-23 | 1.00 | latinx |
| rs35044562 | rs17712877:C | SLC6A20 | breast_epithelium | 1.79E-07 | 0.94 | latinx |
| rs35044562 | rs73064431 | SLC6A20 | esophagus_muscularis_mucosa | 1.47E-08 | 0.89 | latinx |
| rs10504255 | rs9297994 | CYP7A1 | thyroid_gland | 8.06E-09 | 0.97 | latinx |
| rs10504255 | rs8192870 | SDCBP | left_ventricle_myocardium | 9.90E-10 | 0.88 | latinx |
| rs910888 | rs4413223 | KIF3B | renal_tubule | 1.41E-08 | 0.92 | latinx |
| rs910888 | rs1742980 | KIF3B | esophagus_muscularis_mucosa | 5.03E-08 | 0.93 | latinx |
| rs1260326 | rs1260326 | C2orf16 | liver | 4.93E-23 | 1.00 | african american |
| rs35044562 | rs17712877:C | SLC6A20 | breast_epithelium | 1.79E-07 | 0.91 | african american |
| rs10504255 | rs9297994 | CYP7A1 | thyroid_gland | 8.06E-09 | 0.97 | african american |
| rs1260326 | rs4665972 | NRBP1 | renal_tubule | 5.27E-07 | 0.83 | east asian |

| | | | | | | |
|---|---|---|---|---|---|---|
| rs1260326 | rs4665972 | NRBP1 | naive_CD4_T_cell | 2.77E-07 | 0.83 | east asian |
| rs1260326 | rs1260326 | C2orf16 | liver | 4.93E-23 | 1.00 | east asian |
| rs1260326 | rs780096 | FNDC4 | thyroid_carcinoma | 5.26E-10 | 0.95 | east asian |
| rs35044562 | rs17712877:C | SLC6A20 | breast_epithelium | 1.79E-07 | 0.83 | east asian |
| rs35044562 | rs73064431 | SLC6A20 | esophagus_muscularis_mucosa | 1.47E-08 | 0.96 | east asian |
| rs10504255 | rs9297994 | CYP7A1 | thyroid_gland | 8.06E-09 | 0.94 | east asian |
| rs1260326 | rs4665972 | NRBP1 | renal_tubule | 5.27E-07 | 0.85 | south asian |
| rs1260326 | rs4665972 | NRBP1 | naive_CD4_T_cell | 2.77E-07 | 0.85 | south asian |
| rs1260326 | rs1260326 | C2orf16 | liver | 4.93E-23 | 1.00 | south asian |
| rs35044562 | rs17712877:C | SLC6A20 | breast_epithelium | 1.79E-07 | 0.85 | south asian |
| rs35044562 | rs73064431 | SLC6A20 | esophagus_muscularis_mucosa | 1.47E-08 | 0.97 | south asian |
| rs10504255 | rs9297994 | CYP7A1 | thyroid_gland | 8.06E-09 | 0.97 | south asian |
| rs910888 | rs4413223 | KIF3B | renal_tubule | 1.41E-08 | 0.81 | south asian |
| rs910888 | rs1742980 | KIF3B | esophagus_muscularis_mucosa | 5.03E-08 | 0.93 | south asian |

**eqtl** = expression quantitative trait locus; **r square** = LD correlation coefficient

**Supplementary Table 3e: Protein quantitative trait locus signals identified on variant to gene effects for GWAS hits from meta-analysis**

| GWAS SNP | pQTL SNP | pQTL gene | pQTL tissue cell | pQTL P Value | r square between GWAS and pQTL SNP | population |
|---|---|---|---|---|---|---|
| rs1260326 | rs1260326 | GCKR | blood_plasma | 1.35E-31 | 1.00 | european |
| rs1260326 | rs1260326 | GCKR | blood_plasma | 1.35E-31 | 1.00 | latinx |
| rs1260326 | rs1260326 | GCKR | blood_plasma | 1.35E-31 | 1.00 | african american |
| rs1260326 | rs1260326 | GCKR | blood_plasma | 1.35E-31 | 1.00 | east asian |
| rs1260326 | rs1260326 | GCKR | blood_plasma | 1.35E-31 | 1.00 | south asian |

**eqtl** = expression quantitative trait locus; **r square** = LD correlation coefficient

**Supplementary Table 3f: Coding signals identified on variant to gene effects for GWAS hits from meta-analysis**

| GWAS SNP | Coding SNP | GWAS P value | Coding Gene | Consequence | r square between GWAS SNP and Coding SNP | Population |
|---|---|---|---|---|---|---|
| rs1260326 | rs1260326 | 1.98E-08 | GCKR | missense_variant&splice_region_variant | 1 | european |

**r square** = LD correlation coefficient

| Supplementary Table 4: Association of index variants from multi-ancestry meta-analysis with covid measures in 23andMe database | | | | | | | |
|---|---|---|---|---|---|---|---|
| **SNP** | **Phenotype** | **Cases N** | **Controls N** | **Alleles** | **EA(Frequency)** | **OR(95% CI)** | **P value** |
| rs10504255 | COVID-19 Hospitalization | 11874 | 304988 | A/G | A(0.66) | 1.00(0.96,1.03) | 8.63E-01 |
| rs10504255 | COVID-19 test positivity | 334905 | 394557 | A/G | A(0.66) | 1.00(0.99,1.01) | 9.01E-01 |
| rs10504255 | COVID-19 respiratory issues | 19803 | 315102 | A/G | G(0.34) | 1.02(0.99,1.05) | 2.06E-01 |
| rs10504255 | Long COVID | 79816 | 248079 | A/G | G(0.34) | 1.01(0.99,1.03) | 2.73E-01 |
| rs1260326 | COVID-19 Hospitalization | 11874 | 304988 | C/T | T(0.02) | 1.05(0.93.1.19) | 4.37E-01 |
| rs1260326 | COVID-19 test positivity | 334905 | 394557 | C/T | T(0.02) | 0.99(0.96,1.03) | 7.41E-01 |
| rs1260326 | COVID-19 respiratory issues | 19803 | 315102 | C/T | T(0.02) | 1.03(0.94,1.13) | 5.51E-01 |
| rs1260326 | Long COVID | 79816 | 248079 | C/T | T(0.02) | 0.99(0.93,1.05) | 6.40E-01 |
| rs13245319 | COVID-19 Hospitalization | 11874 | 304988 | A/C | A(0.99) | 1.08(0.90,1.29) | 4.11E-01 |
| rs13245319 | COVID-19 test positivity | 334905 | 394557 | A/C | A(0.99) | 0.99(0.95,1.03) | 6.83E-01 |
| rs13245319 | COVID-19 respiratory issues | 19803 | 315102 | A/C | A(0.99) | 0.97(0.85,1.10) | 6.17E-01 |
| rs13245319 | Long COVID | 79816 | 248079 | A/C | A(0.99) | 0.96(0.88,1.04) | 2.98E-01 |
| rs35044562 | COVID-19 Hospitalization | 11874 | 304988 | A/G | G(0.08) | 1.53(1.45,1.62) | 1.10E-56 |
| rs35044562 | COVID-19 test positivity | 334905 | 394557 | A/G | G(0.08) | 1.03(1.02,1.05) | 1.10E-05 |
| rs35044562 | COVID-19 respiratory issues | 19803 | 315102 | A/G | G(0.08) | 1.46(1.40,1.52) | 7.08E-69 |

| | | | | | | | |
|---|---|---|---|---|---|---|---|
| rs35044562 | Long COVID | 79816 | 248079 | A/G | G(0.08) | 1.05(1.02,1.08) | 1.51E-03 |
| rs536843010 | COVID-19 Hospitalization | 11874 | 304988 | A/G | A(0.99) | 1.09(0.35,3.45) | 8.77E-01 |
| rs536843010 | COVID-19 test positivity | 334905 | 394557 | A/G | A(0.99) | 0.99(0.76,1.28) | 9.19E-01 |
| rs536843010 | COVID-19 respiratory issues | 19803 | 315102 | A/G | A(0.99) | 0.79(0.37,1.70) | 5.55E-01 |
| rs536843010 | Long COVID | 79816 | 248079 | A/G | A(0.99) | 1.00(0.59,1.71) | 9.86E-01 |
| rs75683620 | COVID-19 Hospitalization | 11874 | 304988 | C/T | T(0.04) | 1.01(0.97,1.04) | 7.42E-01 |
| rs75683620 | COVID-19 test positivity | 334905 | 394557 | C/T | T(0.04) | 1.00(0.99,1.01) | 7.65E-01 |
| rs75683620 | COVID-19 respiratory issues | 19803 | 315102 | C/T | T(0.04) | 1.02(1.00,1.05) | 8.08E-02 |
| rs75683620 | Long COVID | 79816 | 248079 | C/T | T(0.04) | 1.02(1.01,1.04) | 4.59E-03 |

**EA** = effect allele, **OR** = Odds ratio, **CI** = Confidence Interval

| Supplemental Table 5a: PheWAS of rs10504255 in 23andMe European ancestry population | | | | | | | | | |
|---|---|---|---|---|---|---|---|---|---|
| **SNP** | **Phenotype** | **Alleles** | **Effect allele** | **EAF** | **P value** | **Effect** | **Stderr** | **Control N** | **Case N** |
| rs10504255 | High Cholesterol | A/G | G | 0.344 | 7.53E-172 | 0.073 | 0.0026 | 1471832 | 424454 |
| rs10504255 | IBS Diarrhea | A/G | G | 0.344 | 5.31E-134 | -0.096 | 0.0039 | 3310458 | 157988 |
| rs10504255 | IBS | A/G | G | 0.344 | 3.95E-113 | -0.051 | 0.0023 | 2982083 | 531828 |
| rs10504255 | Gallstone Disease | A/G | G | 0.344 | 3.38E-82 | 0.065 | 0.0034 | 1486267 | 239837 |
| rs10504255 | High LDL | A/G | G | 0.344 | 2.69E-62 | 0.053 | 0.0032 | 1010597 | 284391 |
| rs10504255 | High Triglycerides | A/G | G | 0.344 | 1.47E-60 | 0.056 | 0.0034 | 768258 | 265715 |
| rs10504255 | IBS Mixed type | A/G | G | 0.344 | 1.99E-50 | -0.053 | 0.0036 | 3270426 | 193722 |
| rs10504255 | Cardiometabolic condition <60 years of age | A/G | G | 0.344 | 1.94E-33 | 0.035 | 0.0029 | 353569 | 1154042 |
| rs10504255 | Cardiometabolic condition | A/G | G | 0.344 | 8.06E-32 | 0.010 | 0.0009 | 1189273 | 871097 |
| rs10504255 | Age of high cholesterol | A/G | G | 0.344 | 4.18E-18 | -0.012 | 0.0014 | NA | 1107797 |
| rs10504255 | Diary products indigestions | A/G | G | 0.344 | 4.64E-18 | -0.017 | 0.0020 | 530425 | 158755 |
| rs10504255 | Fatty Liver | A/G | G | 0.344 | 1.35E-15 | 0.032 | 0.0040 | 3449623 | 148386 |
| rs10504255 | IBS Constipation type | A/G | G | 0.344 | 1.74E-14 | 0.047 | 0.0061 | 3416746 | 61548 |
| rs10504255 | Blue eyecolor | A/G | G | 0.344 | 4.58E-14 | -0.008 | 0.0010 | 337491 | 167492 |
| rs10504255 | Fatty Liver and Overweight | A/G | G | 0.344 | 2.22E-13 | 0.037 | 0.0051 | 3516705 | 88060 |
| rs10504255 | Type 2 diabetes | A/G | G | 0.344 | 1.06E-12 | -0.022 | 0.0031 | 3356306 | 284019 |
| rs10504255 | Lactose intolerance | A/G | G | 0.344 | 1.31E-12 | -0.030 | 0.0042 | 781223 | 158772 |
| rs10504255 | High plasma glucose level | A/G | G | 0.344 | 3.58E-11 | -0.016 | 0.0024 | 2999408 | 467867 |
| rs10504255 | High blood pressure | A/G | G | 0.344 | 9.42E-11 | 0.012 | 0.0019 | 2346181 | 1208987 |
| rs10504255 | Female body type | A/G | G | 0.344 | 4.75E-09 | -0.019 | 0.0032 | 501176 | 415015 |

| | | | | | | | | | |
|---|---|---|---|---|---|---|---|---|---|
| rs10504255 | Green eyecolor | A/G | G | 0.344 | 6.69E-09 | -0.009 | 0.0015 | 15483 | 147960 |
| rs10504255 | Height | A/G | G | 0.344 | 2.70E-08 | -0.004 | 0.0008 | NA | 3652184 |
| rs10504255 | Social relationships | A/G | G | 0.344 | 3.37E-08 | 0.006 | 0.0011 | NA | 1693983 |
| rs10504255 | Ever tobacco user | A/G | G | 0.344 | 3.85E-08 | -0.009 | 0.0016 | 1835629 | 1774277 |
| rs10504255 | Early onse of fatty liver (age 35) | A/G | G | 0.344 | 2.27E-07 | 0.045 | 0.0087 | 3523567 | 29175 |
| rs10504255 | Fracture | A/G | G | 0.344 | 3.23E-07 | -0.008 | 0.0016 | 1925340 | 1468031 |
| rs10504255 | Migraine associated with CHD | A/G | G | 0.344 | 6.74E-07 | -0.015 | 0.0029 | 2872915 | 299819 |
| rs10504255 | elevated_liver_test | A/G | G | 0.344 | 9.68E-07 | 0.014 | 0.0028 | 2075206 | 331208 |
| rs10504255 | Statin Dose increase | A/G | G | 0.344 | 2.98E-06 | 0.037 | 0.0079 | 202033 | 42653 |

**Stderr** = standard error, **EAF** = Effect allele frequency

| Supplemental Table 5b: PheWAS of rs35044562 in 23andMe European ancestry population | | | | | | | | | |
|---|---|---|---|---|---|---|---|---|---|
| **SNP** | **Phenotype** | **Alleles** | **Effect allele** | **EAF** | **P value** | **Effect** | **Stderr** | **Control N** | **Case N** |
| rs35044562 | COVID-19+ with sever respiratory disease | A/G | G | 0.078 | 1.11E-68 | 0.405 | 0.0220 | 973358 | 11048 |
| rs35044562 | COVID-19+ with pneumonia | A/G | G | 0.078 | 4.83E-66 | 0.420 | 0.0233 | 974746 | 9739 |
| rs35044562 | COVID-19+ with respiratory support | A/G | G | 0.078 | 1.46E-62 | 0.543 | 0.0305 | 975711 | 5103 |
| rs35044562 | COVID-19+ required hospitalization | A/G | G | 0.078 | 4.59E-54 | 0.452 | 0.0276 | 975366 | 6662 |
| rs35044562 | History of canker sore over past year | A/G | G | 0.078 | 1.55E-13 | -0.054 | 0.0073 | 379246 | 228225 |
| rs35044562 | Hypothyroidism | A/G | G | 0.078 | 3.43E-09 | 0.026 | 0.0045 | 3055549 | 443486 |
| rs35044562 | COVID-19+ test positive vs negative | A/G | G | 0.078 | 1.03E-07 | 0.048 | 0.0090 | 263815 | 151497 |
| rs35044562 | Hashimotos thyroiditis | A/G | G | 0.078 | 4.15E-07 | 0.036 | 0.0070 | 3417912 | 149411 |
| rs35044562 | Celiac disease | A/G | G | 0.078 | 3.18E-06 | 0.089 | 0.0188 | 2101501 | 18977 |
| **Stderr** = standard error, **EAF** = Effect allele frequency | | | | | | | | | |

| Supplemental Table 5c: PheWAS of rs1260326 in 23andMe European ancestry population | | | | | | | | | |
|---|---|---|---|---|---|---|---|---|---|
| SNP | Phenotype | Alleles | Effect allele | EAF | P value | Effect | Stderr | Control N | Case N |
| rs1260326 | Gout | C/T | T | 0.42 | <5.42E-232 | 0.147 | 0.0037 | 3324491 | 170438 |
| rs1260326 | High triglycerides | C/T | T | 0.42 | <5.45E-232 | 0.257 | 0.0033 | 768258 | 265715 |
| rs1260326 | High Cholesterol | C/T | T | 0.42 | <5.45E-232 | 0.149 | 0.0025 | 1471832 | 424454 |
| rs1260326 | Fatty liver | C/T | T | 0.42 | 5.45E-232 | 0.124 | 0.0038 | 3449623 | 148386 |
| rs1260326 | High LDL | C/T | T | 0.42 | 4.05E-209 | 0.094 | 0.0031 | 1010597 | 284391 |
| rs1260326 | Acne | C/T | T | 0.42 | 2.35E-174 | -0.029 | 0.0010 | 280933 | 711725 |
| rs1260326 | Alcohol use over past 2 weeks | C/T | T | 0.42 | 8.07E-172 | -0.037 | 0.0013 | 1219574 | 1087262 |
| rs1260326 | Fatty liver in overweight individuals | C/T | T | 0.42 | 3.08E-170 | 0.136 | 0.0049 | 3516705 | 88060 |
| rs1260326 | HIgh plasma glucose | C/T | T | 0.42 | 3.53E-164 | -0.064 | 0.0024 | 2999408 | 467867 |
| rs1260326 | Current alcohol use | C/T | T | 0.42 | 2.57E-131 | -0.049 | 0.0020 | 2793862 | 627119 |
| rs1260326 | Height | C/T | T | 0.42 | 6.72E-116 | -0.017 | 0.0008 | NA | 3652184 |
| rs1260326 | Type 2 diabetes | C/T | T | 0.42 | 1.33E-112 | -0.058 | 0.0026 | 3045204 | 386880 |
| rs1260326 | Gallstone disease | C/T | T | 0.42 | 1.25E-103 | -0.071 | 0.0033 | 1486267 | 239837 |
| rs1260326 | IBS Diarrhea | C/T | T | 0.42 | 2.93E-101 | 0.079 | 0.0037 | 3310458 | 157988 |
| rs1260326 | IBS | C/T | T | 0.42 | 9.78E-100 | 0.046 | 0.0022 | 2982083 | 531828 |
| rs1260326 | Resting heart rate | C/T | T | 0.42 | 2.69E-98 | 0.024 | 0.0012 | 70120 | 370362 |
| rs1260326 | Gestational diabetes | C/T | T | 0.42 | 1.85E-96 | -0.091 | 0.0044 | 1494007 | 119373 |
| rs1260326 | Kidney Stones | C/T | T | 0.42 | 2.70E-95 | 0.049 | 0.0024 | 2977966 | 434619 |
| rs1260326 | Weight | C/T | T | 0.42 | 1.79E-94 | -0.015 | 0.0008 | NA | 3652184 |
| rs1260326 | Early age of onset ( < 35 years) of fatty liver | C/T | T | 0.42 | 3.79E-86 | 0.165 | 0.0084 | 3523567 | 29175 |
| rs1260326 | Age of graying of hair | C/T | T | 0.42 | 9.42E-83 | 0.077 | 0.0040 | 19977 | 56316 |
| rs1260326 | Severe acne | C/T | T | 0.42 | 1.81E-80 | -0.046 | 0.0024 | 3076780 | 405667 |

| | | | | | | | | | |
|---|---|---|---|---|---|---|---|---|---|
| rs1260326 | Preference to sweet vs salty | C/T | T | 0.42 | 6.44E-75 | -0.052 | 0.0029 | 624616 | 434200 |
| rs1260326 | Cardiometabolic condition | C/T | T | 0.42 | 3.45E-73 | 0.015 | 0.0008 | 1189273 | 871097 |
| rs1260326 | Cardiometabolic condition < 60 years | C/T | T | 0.42 | 2.18E-69 | 0.050 | 0.0028 | 353569 | 1154042 |
| rs1260326 | Sweet tooth | C/T | T | 0.42 | 7.51E-62 | -0.041 | 0.0025 | 1265928 | 464802 |
| rs1260326 | Healthy old | C/T | T | 0.42 | 2.06E-60 | -0.049 | 0.0030 | 2358010 | 255059 |
| rs1260326 | NASH | C/T | T | 0.42 | 2.22E-59 | 0.164 | 0.0101 | 3550649 | 20099 |
| rs1260326 | Obesity | C/T | T | 0.42 | 7.24E-55 | -0.025 | 0.0016 | 2225295 | 1311163 |
| rs1260326 | Number of dental cavities | C/T | T | 0.42 | 1.61E-48 | -0.022 | 0.0015 | 121814 | 306855 |
| rs1260326 | Low HDL | C/T | T | 0.42 | 5.29E-48 | 0.052 | 0.0035 | 847325 | 220173 |
| rs1260326 | Alcohol flush | C/T | T | 0.42 | 1.53E-45 | 0.019 | 0.0013 | 298337 | 145726 |
| rs1260326 | Elevated liver test | C/T | T | 0.42 | 9.39E-39 | 0.035 | 0.0027 | 2075206 | 331208 |
| rs1260326 | IBS Mixed subtype | C/T | T | 0.42 | 1.91E-36 | 0.043 | 0.0034 | 3270426 | 193722 |
| rs1260326 | BMI | C/T | T | 0.42 | 6.79E-35 | -0.009 | 0.0008 | NA | 3652184 |
| rs1260326 | Current caffeine use | C/T | T | 0.42 | 1.70E-33 | -0.006 | 0.0005 | 90209 | 28 |
| rs1260326 | Crohn's disease | C/T | T | 0.42 | 4.91E-31 | 0.086 | 0.0074 | 3557533 | 37969 |
| rs1260326 | Essential tremor | C/T | T | 0.42 | 6.60E-31 | -0.086 | 0.0075 | 3473649 | 38640 |
| rs1260326 | Skin color | C/T | T | 0.42 | 1.76E-30 | -0.015 | 0.0013 | 111675 | 411896 |
| rs1260326 | Age of high cholesterol | C/T | T | 0.42 | 2.94E-30 | -0.015 | 0.0013 | NA | 1107797 |
| rs1260326 | Heavy caffeine drinker | C/T | T | 0.42 | 9.25E-30 | -0.035 | 0.0031 | 518003 | 394996 |
| rs1260326 | Hyperglycemia | C/T | T | 0.42 | 2.65E-27 | -0.041 | 0.0038 | 3510871 | 153880 |
| rs1260326 | Vegetable servings | C/T | T | 0.42 | 1.26E-26 | 0.006 | 0.0006 | 212171 | 2025206 |
| rs1260326 | Eczema | C/T | T | 0.42 | 3.70E-25 | 0.023 | 0.0022 | 2989134 | 504013 |
| rs1260326 | Crohns or ulcerative_colitis | C/T | T | 0.42 | 5.58E-24 | 0.048 | 0.0047 | 3490263 | 93964 |
| rs1260326 | Car sickness | C/T | T | 0.42 | 4.87E-22 | 0.010 | 0.0011 | 856656 | 381741 |
| rs1260326 | low testosterone | C/T | T | 0.42 | 3.13E-21 | 0.062 | 0.0066 | 463571 | 54071 |
| rs1260326 | Headache with red wine | C/T | T | 0.42 | 5.11E-20 | 0.036 | 0.0039 | 606099 | 188802 |
| rs1260326 | Frequency of sugary drinks | C/T | T | 0.42 | 1.07E-19 | -0.012 | 0.0014 | 880180 | 326466 |

| | | | | | | | | | |
|---|---|---|---|---|---|---|---|---|---|
| rs1260326 | Daytime sleepiness | C/T | T | 0.42 | 1.21E-19 | 0.012 | 0.0014 | 171131 | 439789 |
| rs1260326 | Uterine fibroids | C/T | T | 0.42 | 5.37E-19 | 0.026 | 0.0029 | 1883856 | 295020 |
| rs1260326 | Susceptibility to sunburns | C/T | T | 0.42 | 9.81E-19 | -0.014 | 0.0015 | 88543 | 178389 |
| rs1260326 | HIgh blood pressure | C/T | T | 0.42 | 4.76E-18 | 0.016 | 0.0018 | 2346181 | 1208987 |
| rs1260326 | Sunburns | C/T | T | 0.42 | 3.69E-17 | 0.028 | 0.0034 | 466977 | 322704 |
| rs1260326 | History of blushing easily | C/T | T | 0.42 | 3.85E-17 | 0.025 | 0.0030 | 575609 | 460048 |
| rs1260326 | Likeness to vanilla ice cream flavor | C/T | T | 0.42 | 6.24E-17 | -0.020 | 0.0024 | 897369 | 640185 |
| rs1260326 | Diagnosis of migraine | C/T | T | 0.42 | 6.38E-17 | 0.016 | 0.0019 | 2654329 | 808027 |
| rs1260326 | HIP Allergies | C/T | T | 0.42 | 9.41E-17 | 0.013 | 0.0016 | 1950163 | 1505262 |
| rs1260326 | Any allergy | C/T | T | 0.42 | 9.45E-17 | 0.013 | 0.0016 | 1969024 | 1522882 |
| rs1260326 | Osteoporosis | C/T | T | 0.42 | 1.33E-16 | 0.024 | 0.0029 | 3062950 | 394378 |
| rs1260326 | education over 30 years | C/T | T | 0.42 | 1.86E-16 | 0.017 | 0.0021 | 33408 | 332373 |
| rs1260326 | Abnormal liver test | C/T | T | 0.42 | 3.71E-16 | 0.047 | 0.0058 | 753690 | 67499 |
| rs1260326 | Psoriasis | C/T | T | 0.42 | 4.33E-16 | 0.026 | 0.0032 | 3309074 | 216644 |
| rs1260326 | Diabetic neuropathy | C/T | T | 0.42 | 1.17E-15 | 0.056 | 0.0069 | 220464 | 54686 |
| rs1260326 | On anti-tnf alpha medications | C/T | T | 0.42 | 1.55E-15 | 0.048 | 0.0060 | 3509963 | 58259 |
| rs1260326 | Frequency of green vegetables intake | C/T | T | 0.42 | 3.28E-15 | 0.008 | 0.0010 | 78516 | 447720 |
| rs1260326 | Elevated AST | C/T | T | 0.42 | 4.78E-15 | 0.054 | 0.0068 | 438164 | 49154 |
| rs1260326 | Age at menopause | C/T | T | 0.42 | 6.34E-15 | 0.021 | 0.0027 | 11549 | 9929 |
| rs1260326 | First born birth weight | C/T | T | 0.42 | 9.11E-15 | -0.016 | 0.0020 | 8430 | 7293 |
| rs1260326 | Frequency of green leafy vegetables | C/T | T | 0.42 | 1.49E-14 | 0.010 | 0.0013 | 126291 | 672486 |
| rs1260326 | Male pattern of baldness | C/T | T | 0.42 | 1.55E-14 | -0.028 | 0.0036 | 307522 | 426179 |
| rs1260326 | Liver fibrosis | C/T | T | 0.42 | 1.64E-14 | 0.210 | 0.0273 | 520224 | 2735 |
| rs1260326 | Nearsightedness | C/T | T | 0.42 | 6.08E-14 | 0.018 | 0.0024 | 679860 | 869024 |
| rs1260326 | Liking for spicy food | C/T | T | 0.42 | 9.76E-14 | 0.015 | 0.0020 | 156168 | 176177 |

| | | | | | | | | | |
|---|---|---|---|---|---|---|---|---|---|
| rs1260326 | Preference of chocolate ice cream | C/T | T | 0.42 | 1.01E-13 | 0.017 | 0.0023 | 890148 | 669470 |
| rs1260326 | Age at high blood pressure | C/T | T | 0.42 | 2.17E-13 | -0.010 | 0.0014 | NA | 1078184 |
| rs1260326 | Microvascular conditions | C/T | T | 0.42 | 2.41E-13 | 0.046 | 0.0062 | 200012 | 74996 |
| rs1260326 | Increase in statin dose | C/T | T | 0.42 | 9.48E-13 | 0.054 | 0.0076 | 202033 | 42653 |
| rs1260326 | Hair pattern - curly | C/T | T | 0.42 | 1.48E-12 | -0.012 | 0.0017 | 301345 | 333924 |
| rs1260326 | History of nosebleeds | C/T | T | 0.42 | 1.57E-12 | 0.040 | 0.0056 | 893266 | 71540 |
| rs1260326 | Morning person | C/T | T | 0.42 | 6.19E-12 | -0.013 | 0.0018 | 1351326 | 1354530 |
| rs1260326 | Age of first menses | C/T | T | 0.42 | 6.98E-12 | 0.009 | 0.0013 | 1109 | 23731 |
| rs1260326 | History of pollen allergy | C/T | T | 0.42 | 7.59E-12 | 0.016 | 0.0024 | 2265190 | 462320 |
| rs1260326 | History of seasonal allergies | C/T | T | 0.42 | 9.88E-12 | 0.015 | 0.0022 | 2233928 | 530564 |
| rs1260326 | History of food allergies | C/T | T | 0.42 | 1.07E-11 | 0.023 | 0.0033 | 2388594 | 207577 |
| rs1260326 | Rhinitis | C/T | T | 0.42 | 2.23E-11 | 0.015 | 0.0023 | 2246185 | 522069 |
| rs1260326 | History of animal allergies | C/T | T | 0.42 | 4.48E-11 | 0.018 | 0.0027 | 2327790 | 322081 |
| rs1260326 | History of plant allergies | C/T | T | 0.42 | 4.62E-11 | 0.015 | 0.0023 | 2254538 | 485038 |
| rs1260326 | Skin tags | C/T | T | 0.42 | 5.31E-11 | -0.019 | 0.0029 | 531231 | 474532 |
| rs1260326 | History of stretch marks | C/T | T | 0.42 | 7.95E-11 | -0.019 | 0.0030 | 651955 | 528850 |
| rs1260326 | History of neuropathy | C/T | T | 0.42 | 8.39E-11 | 0.012 | 0.0018 | 390722 | 248221 |
| rs1260326 | Dry skins | C/T | T | 0.42 | 8.45E-11 | 0.009 | 0.0015 | 170649 | 297173 |
| rs1260326 | Food allergy requiring epipen | C/T | T | 0.42 | 2.88E-10 | 0.041 | 0.0065 | 643059 | 53882 |
| rs1260326 | Freckling | C/T | T | 0.42 | 3.22E-10 | 0.008 | 0.0013 | 210274 | 370085 |
| rs1260326 | Allergy to weeds | C/T | T | 0.42 | 3.70E-10 | 0.017 | 0.0027 | 2315486 | 350333 |
| rs1260326 | Statin side-effect on liver function | C/T | T | 0.42 | 3.91E-10 | 0.074 | 0.0119 | 335784 | 15209 |
| rs1260326 | Canker sore | C/T | T | 0.42 | 7.61E-10 | 0.021 | 0.0034 | 249600 | 681087 |
| rs1260326 | Snoring | C/T | T | 0.42 | 9.28E-10 | -0.019 | 0.0030 | 567427 | 399560 |
| rs1260326 | Juvenile asthma | C/T | T | 0.42 | 1.26E-09 | 0.014 | 0.0023 | 2830291 | 472162 |

| | | | | | | | | | |
|---|---|---|---|---|---|---|---|---|---|
| rs1260326 | Tonsillectomy | C/T | T | 0.42 | 1.39E-09 | 0.015 | 0.0024 | 1100056 | 614894 |
| rs1260326 | Emotional eating | C/T | T | 0.42 | 1.85E-09 | 0.033 | 0.0056 | 176017 | 135681 |
| rs1260326 | Coronary artery disease | C/T | T | 0.42 | 3.22E-09 | 0.024 | 0.0041 | 3481135 | 145438 |
| rs1260326 | Age of asthma | C/T | T | 0.42 | 3.77E-09 | -0.010 | 0.0017 | NA | 726853 |
| rs1260326 | Myopia | C/T | T | 0.42 | 8.37E-09 | -0.020 | 0.0034 | 811871 | 263710 |
| rs1260326 | Cold sores | C/T | T | 0.42 | 1.00E-08 | 0.017 | 0.0029 | 532280 | 451022 |
| rs1260326 | Dry eyes | C/T | T | 0.42 | 1.46E-08 | 0.013 | 0.0023 | 2913498 | 514176 |
| rs1260326 | Itchy skin | C/T | T | 0.42 | 2.67E-08 | 0.008 | 0.0014 | 68367 | 283920 |
| rs1260326 | Current tobacco user | C/T | T | 0.42 | 5.02E-08 | -0.012 | 0.0022 | 3167601 | 518972 |
| rs1260326 | History of numbness in extremities | C/T | T | 0.42 | 5.14E-08 | 0.017 | 0.0031 | 530139 | 388175 |
| rs1260326 | Elevated AST | C/T | T | 0.42 | 5.86E-08 | 0.038 | 0.0069 | 431611 | 48347 |
| rs1260326 | History of osteoarthritis of hand vs hip | C/T | T | 0.42 | 6.02E-08 | 0.052 | 0.0096 | 75767 | 33642 |
| rs1260326 | Eating disorder | C/T | T | 0.42 | 8.37E-08 | 0.021 | 0.0040 | 3373897 | 142889 |
| rs1260326 | osteoarthritis | C/T | T | 0.42 | 9.33E-08 | -0.012 | 0.0022 | 2735488 | 634058 |
| rs1260326 | iqb.pastry_frequency | C/T | T | 0.42 | 1.11E-07 | -0.009 | 0.0017 | 242987 | 429825 |
| rs1260326 | Crohn's disease with stricture | C/T | T | 0.42 | 1.30E-07 | 0.097 | 0.0183 | 1100949 | 6190 |
| rs1260326 | Chronic fatigue | C/T | T | 0.42 | 1.73E-07 | 0.021 | 0.0041 | 3357900 | 129306 |
| rs1260326 | Pediatric IBD | C/T | T | 0.42 | 1.75E-07 | 0.065 | 0.0124 | 3539479 | 13347 |
| rs1260326 | Restless leg syndrome | C/T | T | 0.42 | 2.01E-07 | 0.016 | 0.0030 | 3214971 | 251041 |
| rs1260326 | Hepatitis A | C/T | T | 0.42 | 2.15E-07 | 0.042 | 0.0081 | 3572146 | 32139 |
| rs1260326 | Ulcerative Colitis | C/T | T | 0.42 | 2.35E-07 | 0.029 | 0.0057 | 3523877 | 65745 |
| rs1260326 | History of yeast infections | C/T | T | 0.42 | 3.16E-07 | -0.009 | 0.0018 | 127383 | 215379 |
| rs1260326 | Allergy to dust mites | C/T | T | 0.42 | 3.27E-07 | 0.014 | 0.0028 | 2329662 | 318820 |
| rs1260326 | History of higher doses of pain medication | C/T | T | 0.42 | 4.24E-07 | -0.014 | 0.0028 | 1113889 | 364943 |
| rs1260326 | History of carrying epipen | C/T | T | 0.42 | 5.69E-07 | 0.020 | 0.0039 | 1358650 | 149792 |

| | | | | | | | | | |
|---|---|---|---|---|---|---|---|---|---|
| rs1260326 | Melanoma | C/T | T | 0.42 | 6.66E-07 | 0.022 | 0.0044 | 3432878 | 112901 |
| rs1260326 | Immunodeficiency | C/T | T | 0.42 | 6.80E-07 | 0.021 | 0.0043 | 3392818 | 117323 |
| rs1260326 | Squamous cell carcinoma | C/T | T | 0.42 | 7.45E-07 | 0.018 | 0.0035 | 3328414 | 196382 |
| rs1260326 | Dairy products indigestion | C/T | T | 0.42 | 7.79E-07 | 0.009 | 0.0019 | 530425 | 158755 |
| rs1260326 | Binge eating | C/T | T | 0.42 | 1.26E-06 | 0.009 | 0.0018 | 227233 | 192896 |
| rs1260326 | Sweaty palms | C/T | T | 0.42 | 1.35E-06 | 0.023 | 0.0049 | 853183 | 106867 |
| rs1260326 | Frequency of breakfast | C/T | T | 0.42 | 1.46E-06 | -0.010 | 0.0020 | 109644 | 176036 |
| rs1260326 | Ulcer | C/T | T | 0.42 | 1.90E-06 | 0.016 | 0.0033 | 3280680 | 202802 |
| rs1260326 | History of strept throat | C/T | T | 0.42 | 1.96E-06 | 0.007 | 0.0015 | 328700 | 359989 |
| rs1260326 | Psoriatic arthritis | C/T | T | 0.42 | 2.03E-06 | 0.042 | 0.0089 | 3567312 | 26148 |
| rs1260326 | Freckling | C/T | T | 0.42 | 2.36E-06 | 0.017 | 0.0036 | 627439 | 228089 |
| rs1260326 | Severe psoriasis | C/T | T | 0.42 | 2.39E-06 | 0.039 | 0.0084 | 3415996 | 29782 |
| rs1260326 | Allergy to wheat | C/T | T | 0.42 | 2.59E-06 | 0.038 | 0.0082 | 2456856 | 31674 |
| rs1260326 | Rheumatoid arthritis | C/T | T | 0.42 | 3.02E-06 | 0.018 | 0.0039 | 3418053 | 145783 |
| rs1260326 | Blood clots | C/T | T | 0.42 | 4.64E-06 | -0.018 | 0.0039 | 3431131 | 145619 |
| rs1260326 | Side effect of statin use | C/T | T | 0.42 | 5.41E-06 | 0.023 | 0.0050 | 222339 | 128282 |
| rs1260326 | Chronic hives | C/T | T | 0.42 | 5.58E-06 | 0.012 | 0.0026 | 3116460 | 346246 |

**Stderr** = standard error, **EAF** = Effect allele frequency

| Supplemental Table 5d: PheWAS of rs13245319 in 23andME European ancestry population | | | | | | | | | |
|---|---|---|---|---|---|---|---|---|---|
| **SNP** | **Phenotype** | **Alleles** | **Effect allele** | **EAF** | **pvalue** | **effect** | **stderr** | **Control N** | **Case N** |
| rs13245319 | History of kidney stones | C/T | T | 0.021 | 7.84E-08 | 0.047 | 0.0088 | 2977966 | 434619 |
| rs13245319 | IBS | C/T | T | 0.021 | 8.11E-07 | 0.040 | 0.0081 | 2982083 | 531828 |
| rs13245319 | Obesity | C/T | T | 0.021 | 1.25E-06 | -0.029 | 0.0060 | 2225295 | 1311163 |
| rs13245319 | Frequency of back pain | C/T | T | 0.021 | 2.04E-06 | 0.022 | 0.0045 | 613246 | 721643 |
| **Stderr** = standard error, **EAF** = Effect allele frequency | | | | | | | | | |

| Supplementary Table 6: MR Egger slope intercepts representing directional pleiotropy | | | |
|---|---|---|---|
| **Exposure** | **Outcome** | **MR Egger Slope Intercept B(Stderr)** | **P value** |
| IBS -C | covid_symptoms_diarrhea | 0.008(0.008) | 0.3180657654 |
| IBS -D | covid_symptoms_diarrhea | -0.012(0.002) | 1.74E-07 |
| **B** = Intercept estimate, **Stderr** = standard error | | | |

**Supplementary Table 7: Comparison of MR estimates between overlapping and non-overlapping samples**

| Exposure | | Overlapping sample | | | Non-overlapping sample | | |
|---|---|---|---|---|---|---|---|
| | Method | nSNPs | meanF | OR(95%CI) | nSNPs | meanF | OR(95%CI) |
| IBS-D | RE-IVW | 180 | 50.24 | 1.40(1.33, 1.47) | 155 | 50.4 | 1.41(1.33,1.48) |
| | Steiger filtered | 177 | 50.48 | 1.39(1.32, 1.46) | 153 | 50.6 | 1.40(1.33,1.47) |
| | Outlier filtered | 160 | 45.25 | 1.36(1.30, 1.42) | 137 | 45.1 | 1.39(1.33,1.46) |
| IBS-C | RE-IVW | 52 | 43.59 | 0.86(0.79, 0.94) | 50 | 44.1 | 0.86(0.79, 0.94) |
| | Steiger filtered | 51 | 43.18 | 0.89(0.82, 0.96) | 49 | 43.6 | 0.89(0.82, 0.96) |
| | Outlier filtered | 43 | 41.2 | 0.86 (0.81, 0.91) | 41 | 41.8 | 0.86(0.81, 0.91) |

IBS-D = IBS diarrhea, IBS-C = IBS constipation, RE-IVW = random effects inverse variance weighted, nSNPs = number of SNPs, meanF = Mean F statistics, OR = Odds ratio, CI = Confidence Interval

**Supplementary Figure 1: Regional plot around *CYP7A1* Locus for COVID-19+ diarrhea.**
The colors indicate the strength of LD relative to the index variant (rs10504255). The index variant is represented by gray color. Imputed variants are indicated with '+' symbols or '×' symbols for coding variants. Directly genotyped variants are indicated by 'o' symbols or diamond symbols for coding variants.

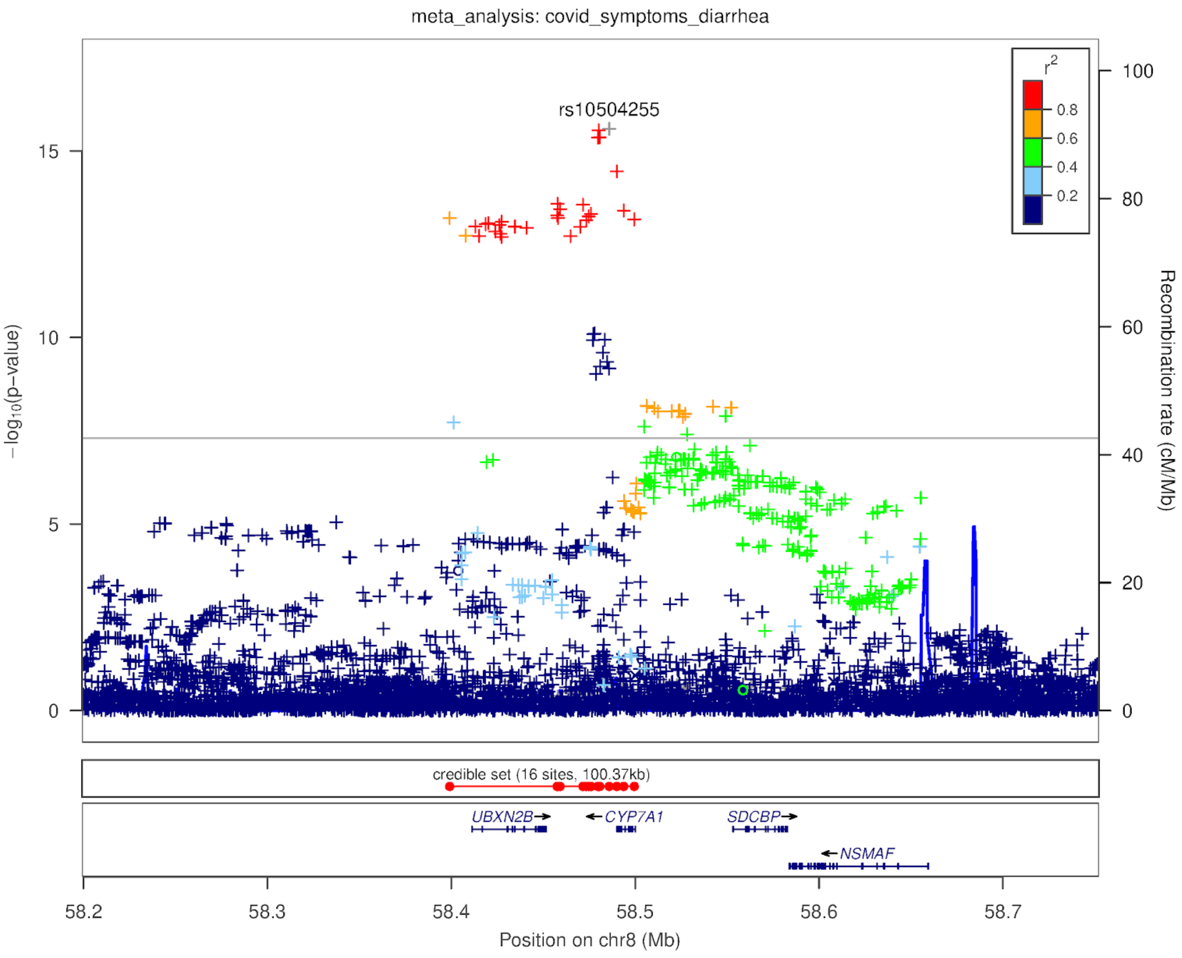

**Supplementary Figure 2: Regional plot around *LZFTL1–CCR9* Locus for COVID-19+ diarrhea.**

The colors indicate the strength of LD relative to the index variant (rs35044562). The index variant is represented by gray color. Imputed variants are indicated with '+' symbols or '×' symbols for coding variants. Directly genotyped variants are indicated by 'o' symbols or diamond symbols for coding variants

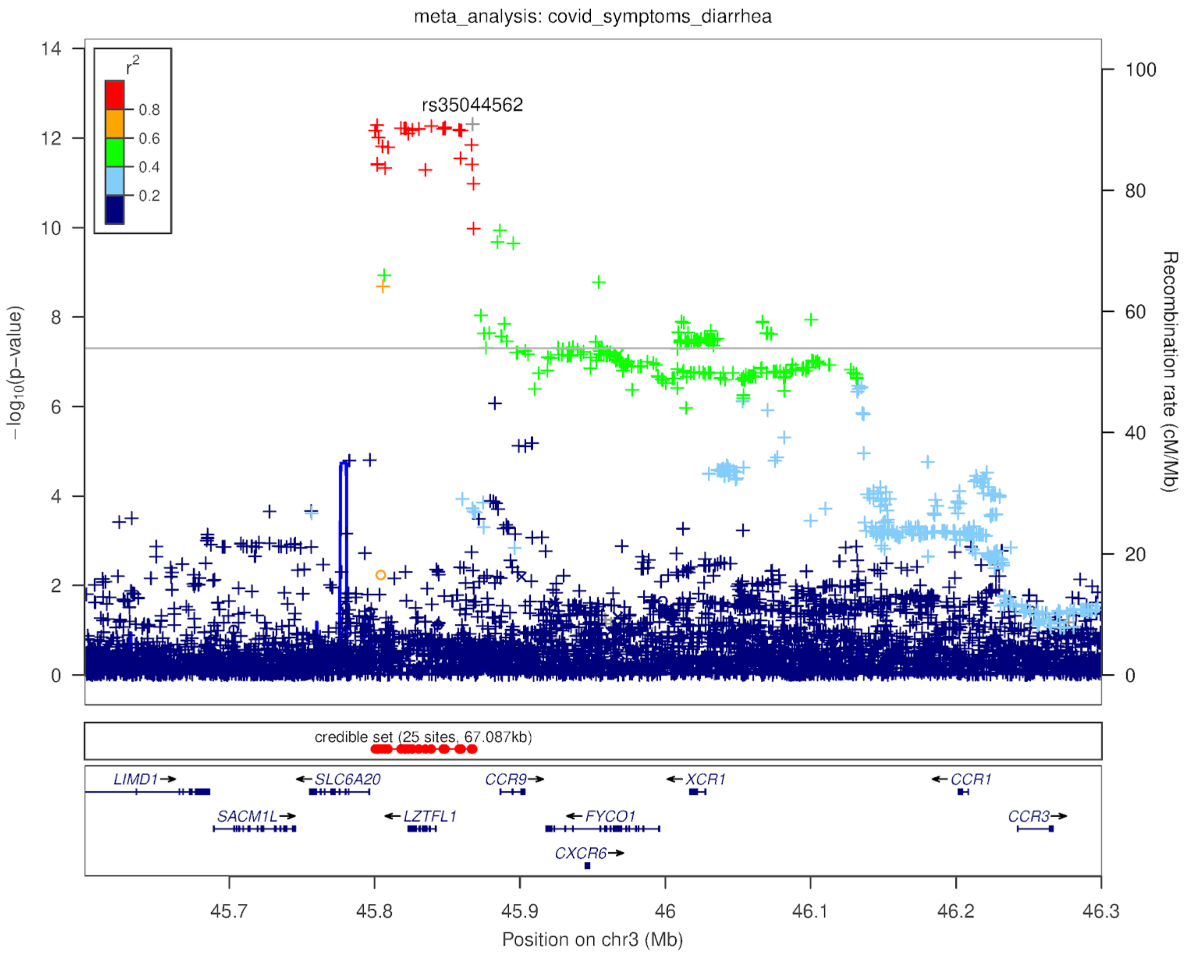

**Supplementary Figure 3: Regional plot around *TMEM182* Locus for COVID-19+ diarrhea.**

The colors indicate the strength of LD relative to the index variant (rs35044562). The index variant is represented by gray color. Imputed variants are indicated with '+' symbols or '×' symbols for coding variants. Directly genotyped variants are indicated by 'o' symbols or diamond symbols for coding variants.

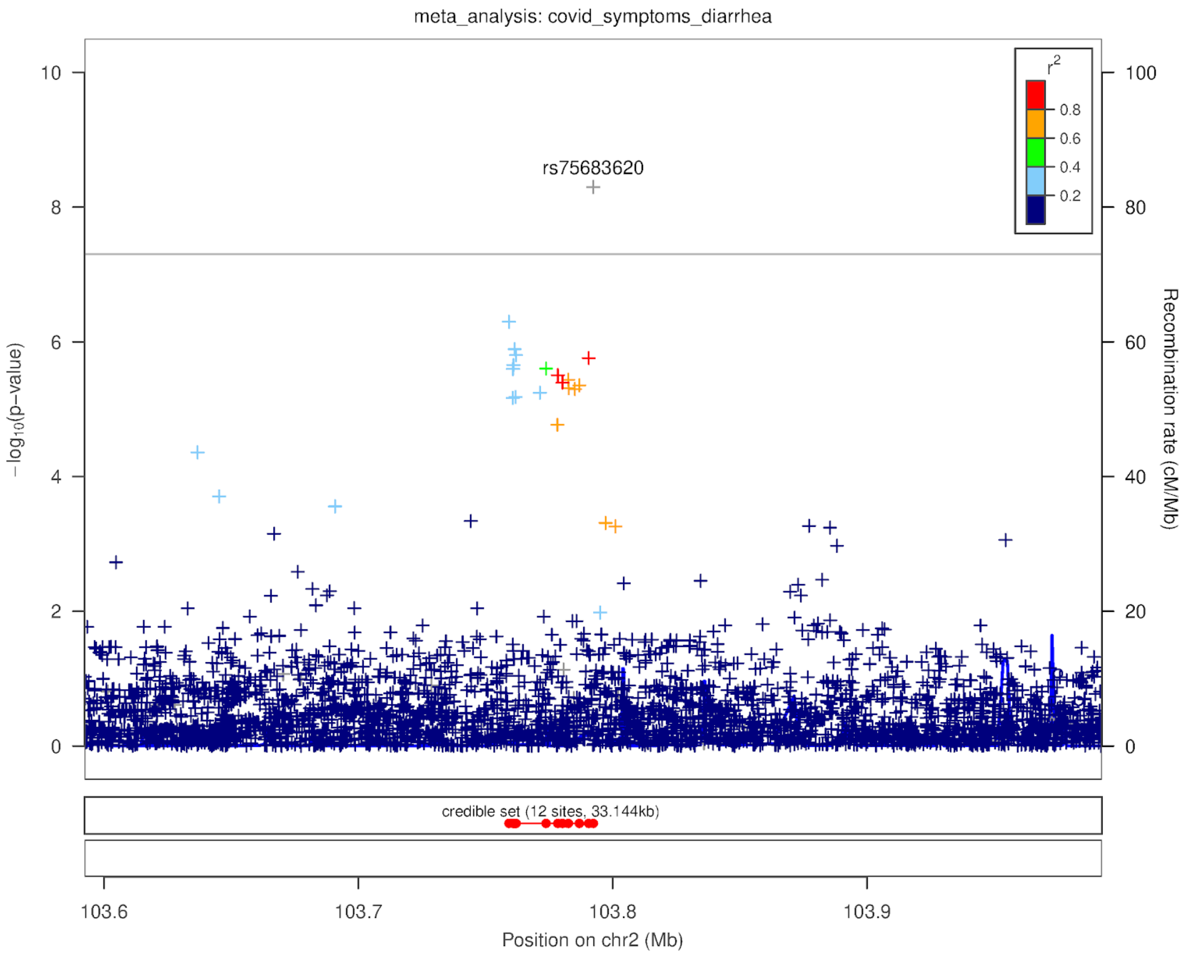

**Supplementary Figure 4: Regional plot around *NALCN* Locus for COVID-19+ diarrhea.**

The colors indicate the strength of LD relative to the index variant (rs536843010). The index variant is represented by gray color. Imputed variants are indicated with '+' symbols or '×' symbols for coding variants. Directly genotyped variants are indicated by 'o' symbols or diamond symbols for coding variants.

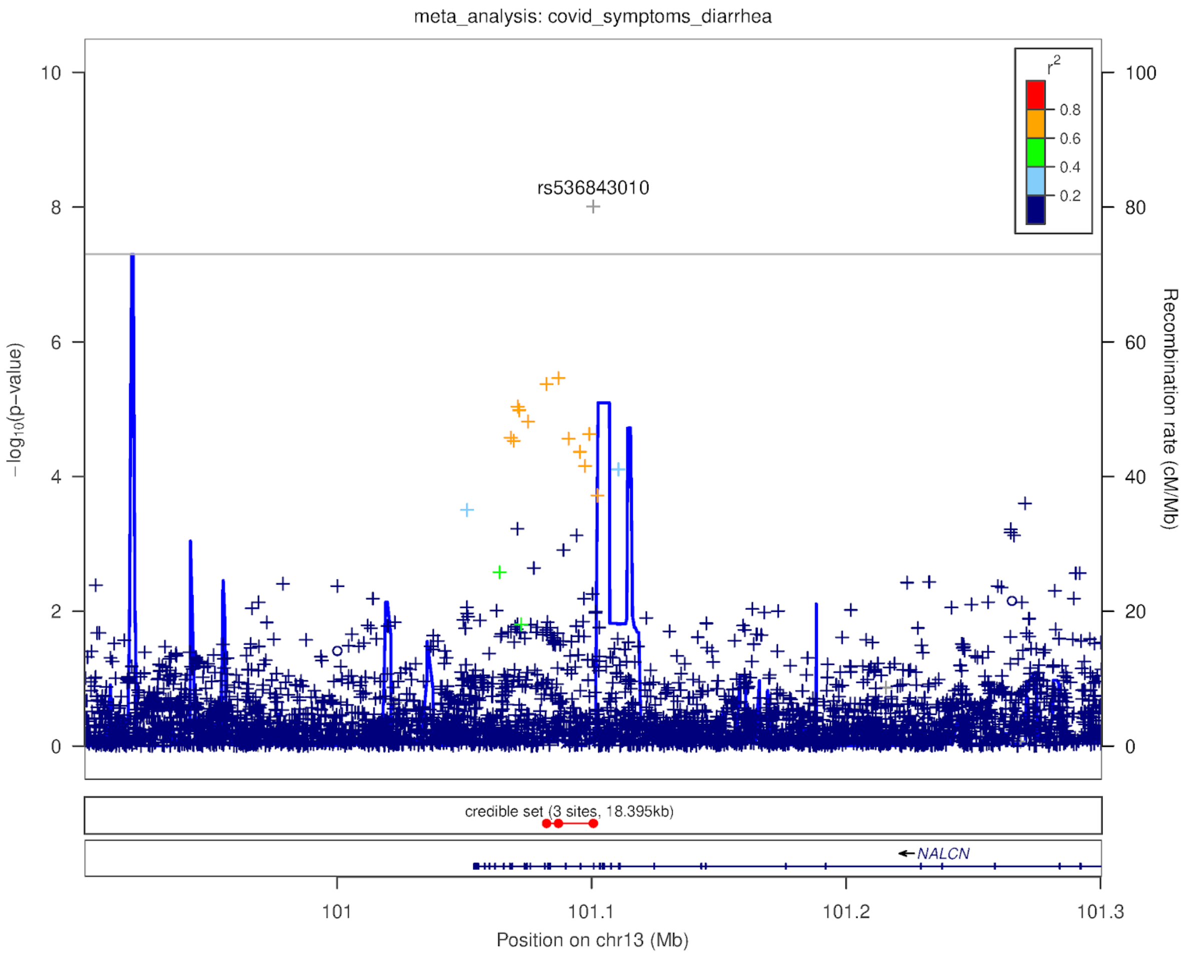

**Supplementary Figure 5: Regional plot around *LFNG* locus for COVID-19+ diarrhea.**

The colors indicate the strength of LD relative to the index variant (rs13245319). The index variant is represented by gray color. Imputed variants are indicated with '+' symbols or '×' symbols for coding variants. Directly genotyped variants are indicated by 'o' symbols or diamond symbols for coding variants.

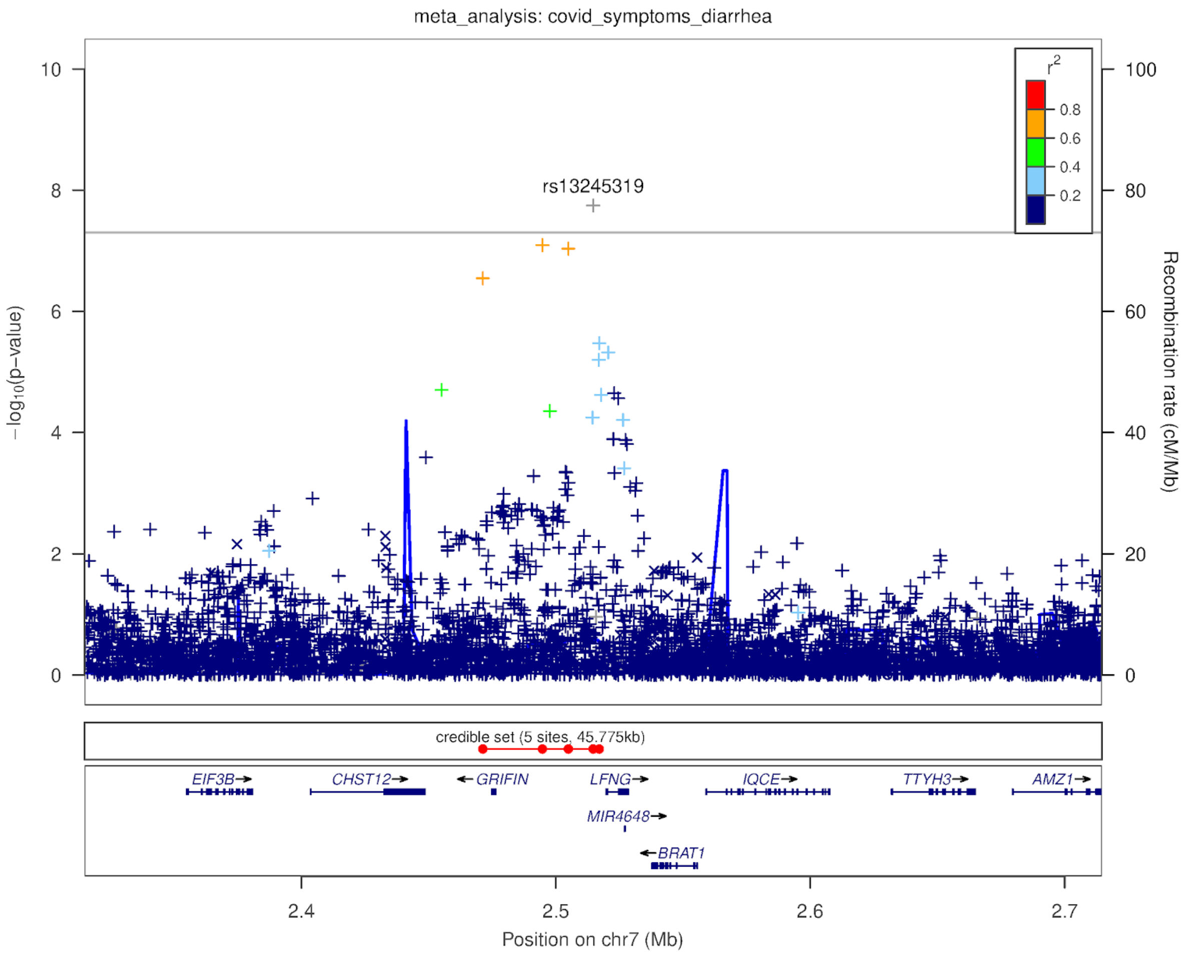

**Supplementary Figure 6: Regional plot around *GCKR* Locus for COVID-19+ diarrhea.**

The colors indicate the strength of LD relative to the index variant (rs1260326). The index variant is represented by gray color. Imputed variants are indicated with '+' symbols or '×' symbols for coding variants. Directly genotyped variants are indicated by 'o' symbols or diamond symbols for coding variants.

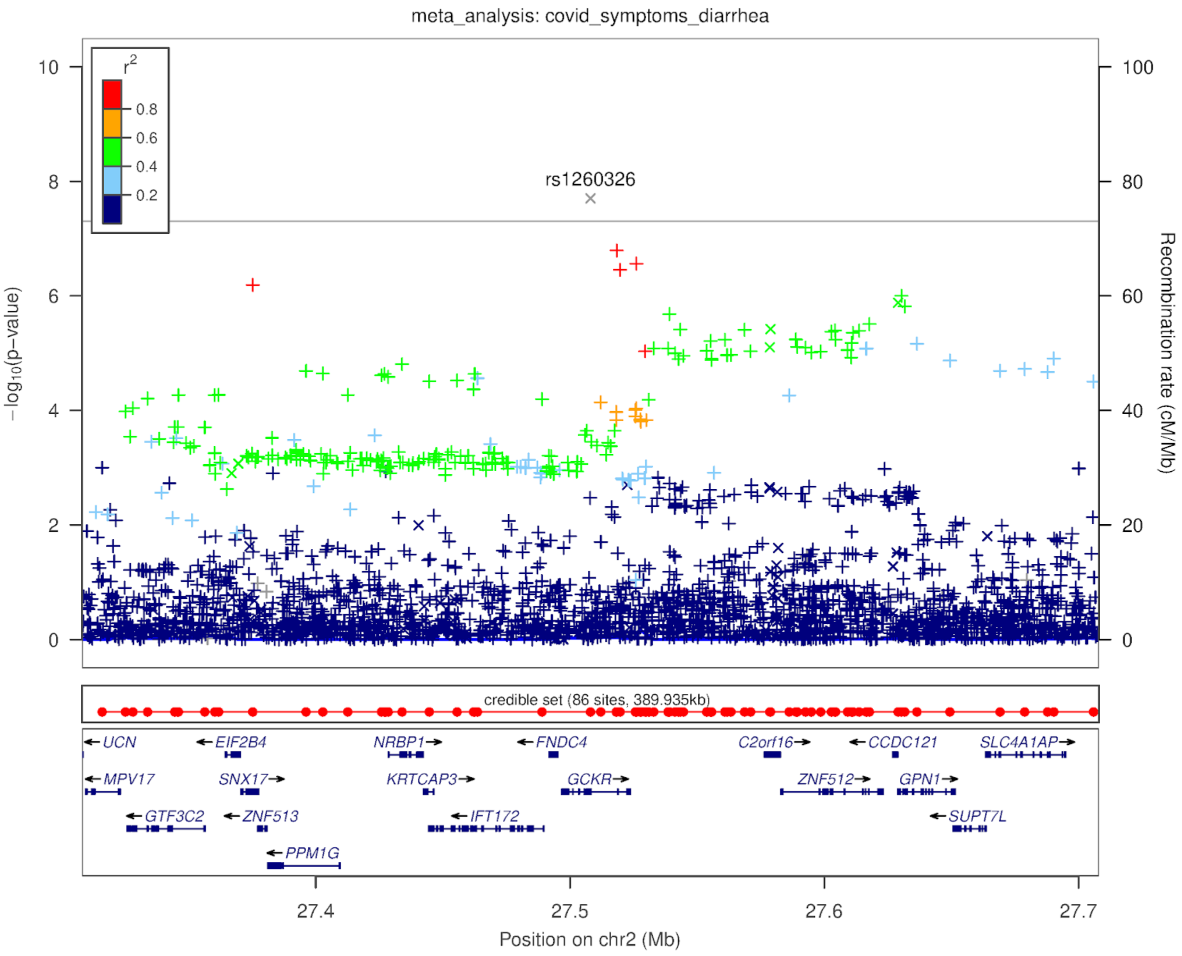

**Supplementary Figure 7**: **Association of GWAS significant hits of COVID-19+ diarrhea with IBS and IBS subtypes in 23andMe participants of European ancestry**

Forest plot represents association of six loci with IBS and IBS subtypes. The results are presented as odds ratio with 95% confidence interval under additive model for allele of each loci to represent increased odds of having COVID-19+ diarrhea. X-axis shows estimates on log scale. Y-axis shows phenotypes studied.

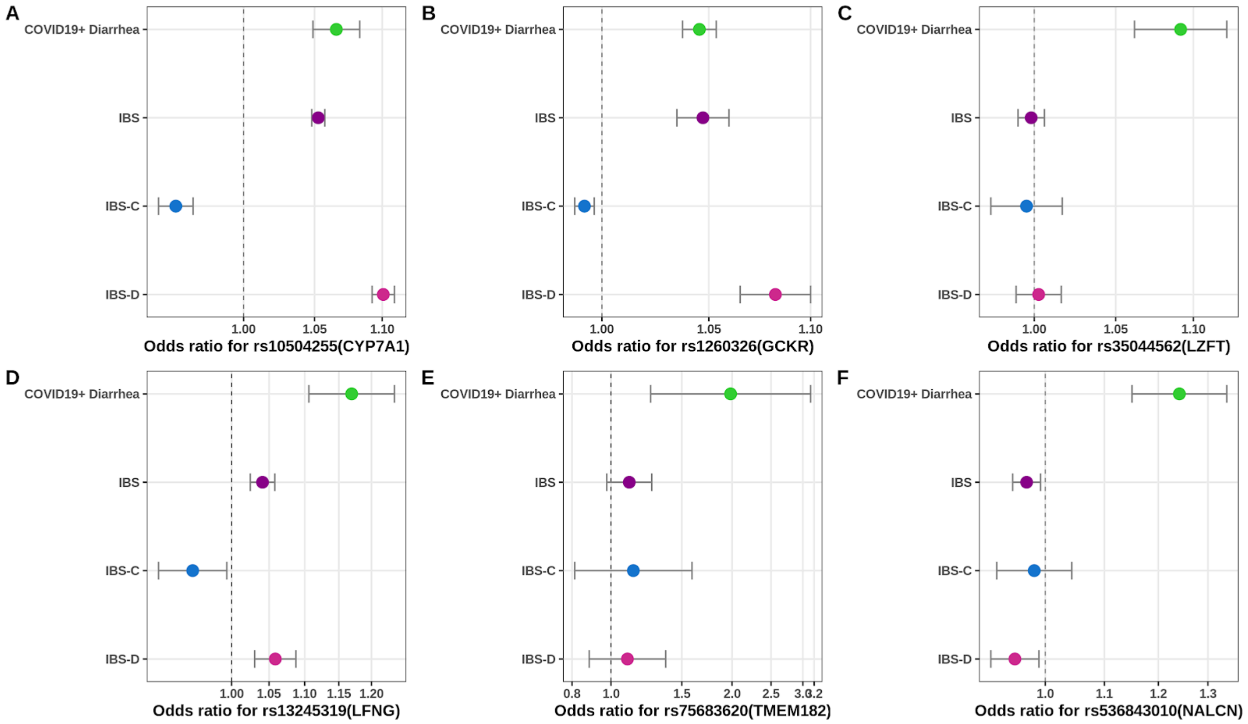

**Supplementary Figure 8: MR scatterplot of genetic associations between IBS subtypes and COVID-19+ diarrhea**

Left side plot**:** The genetic association and corresponding 95% confidence interval (CI) for each SNP (n = 52) with IBS-C (x-axis) and COVID-19+ diarrhea (y-axis) are plotted. The plot represents estimates from three methods: random-effects inverse weighted variance method, weighted median, and random-effects MR Egger.

Right side plot**:** The genetic association and corresponding 95% confidence interval (CI) for each SNP (n = 180) with IBS-D (x-axis) and COVID-19+ diarrhea (y-axis) are plotted. The plot represents estimates from three methods: random-effects inverse weighted variance method, weighted median, and random-effects MR Egger.

The MR Egger intercepts are provided in Supplementary Table 6.

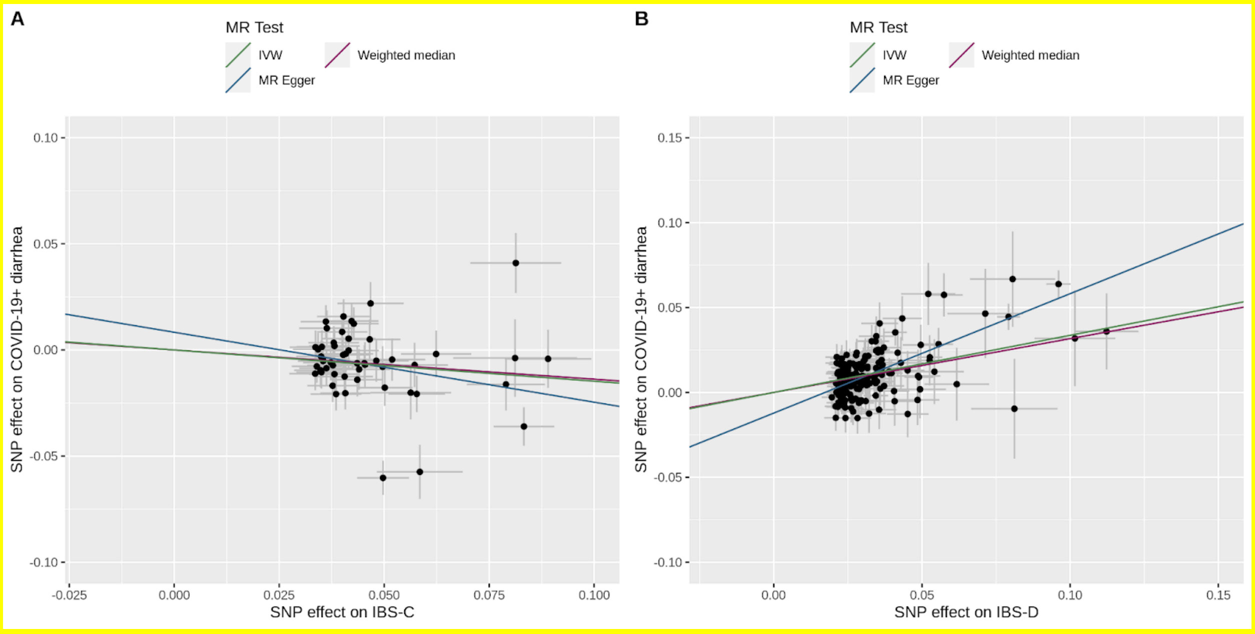

**Supplementary Figure 9: Venn diagram representing overlap between samples used in GWAS of IBS subtypes and COVID-19+ diarrhea in European ancestry**

Panel A: Represents the sample overlap for IBS-D and COVID-19+ diarrhea

Panel B: Represents the sample overlap for IBS-C and COVID-19+ diarrhea

Panel C: Represents overlap between IBS-D cases and COVID-19+ diarrhea cases

Panel D: Represents overlap between IBS-C cases and COVID-19+ diarrhea cases

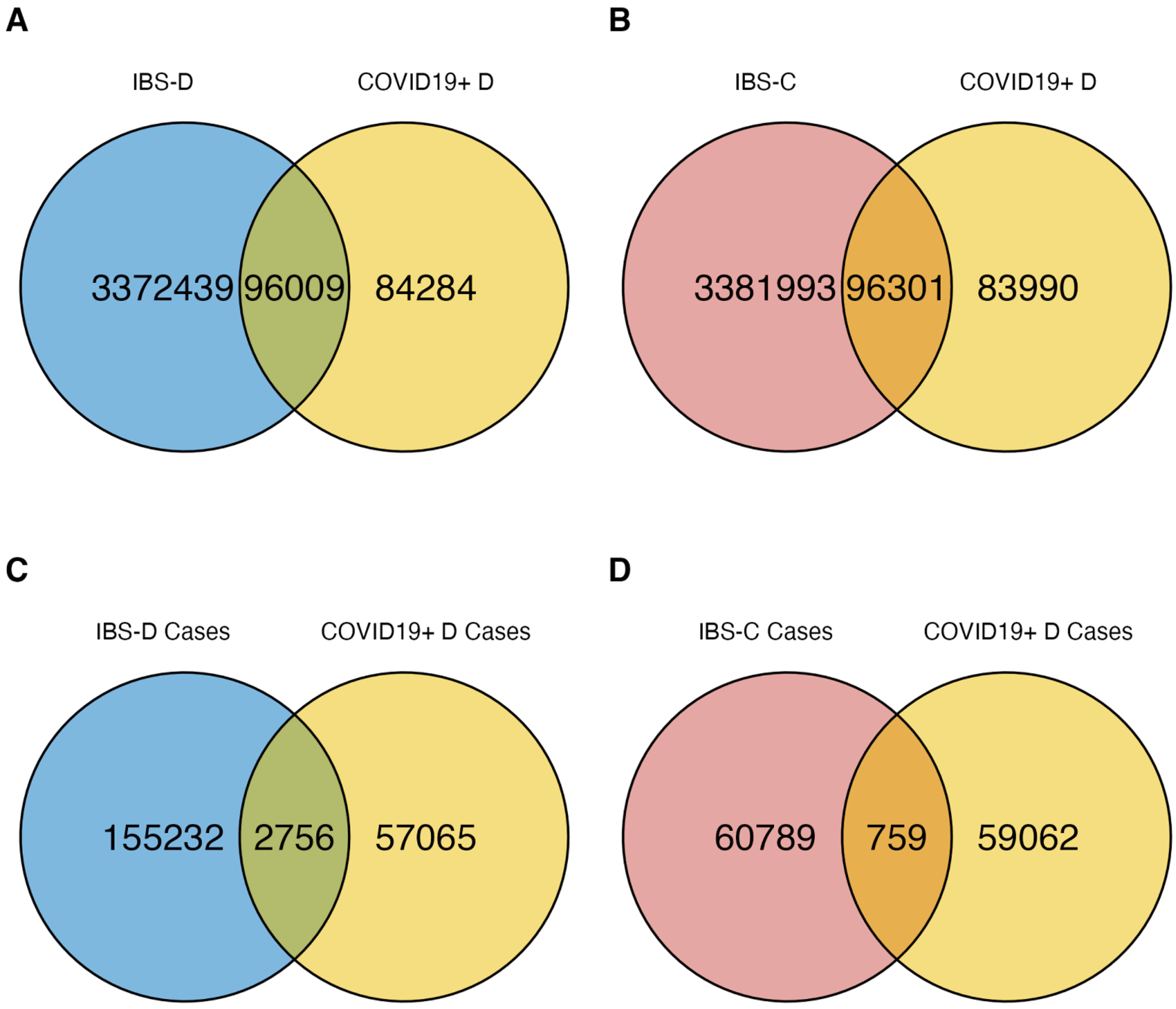
