## Supplementary Data File for "Multi-ancestry GWAS of diarrhea during acute SARS-CoV2 infection identifies multiple novel loci and contrasting etiological roles of irritable bowel syndrome subtypes"

| **Supplementary File Table 1**: **Public datasets that are internally processed using 23andme first eQTL pipeline** | | | | |
| --- | --- | --- | --- | --- |
| **Title** | **PMID** | **Tissue** | **Ethnicity** | **Sample** |
| Transcriptome and genome sequencing uncovers functional variation in humans | [24037378](https://pubmed.ncbi.nlm.nih.gov/24037378/) | Lymphoblast | European | 358 |
| RNA sequencing of whole blood reveals early alterations in immune cells and gene expression in Parkinson's disease | [37117765](https://pubmed.ncbi.nlm.nih.gov/37117765/) | Venous blood | Multi-ethnic | 1,305 |
| The GTEx Consortium atlas of genetic regulatory effects across human tissues | [32913098](https://pubmed.ncbi.nlm.nih.gov/32913098/) | Multiple tissues | Multi-ethnic | 948 |

| **Supplemental File Table 2**: **Public datasets that are internally processed using 23andme second eQTL pipeline** | | | | |
| --- | --- | --- | --- | --- |
| **Title** | **PMID** | **Tissue** | **Ethnicity** | **Sample** |
| Transcriptome and genome sequencing uncovers functional variation in humans | [24037378](https://pubmed.ncbi.nlm.nih.gov/24037378/) | Lymphoblast | European | 358 |
| Characterizing the genetic basis of transcriptome diversity through RNA-sequencing of 922 individuals | [24092820](https://pubmed.ncbi.nlm.nih.gov/2409280/) | Venous blood | NA | 922 |
| RNA sequencing of whole blood reveals early alterations in immune cells and gene expression in Parkinson's disease | [37117765](https://pubmed.ncbi.nlm.nih.gov/37117765/) | Venous blood | European | 752 |
